# Identification of novel plasma lipid markers of cardiovascular disease risk in White and Black women

**DOI:** 10.1101/2022.08.24.22279186

**Authors:** Raghav Jain, Jessica Davidson, Paula Gonzalez, Chris Coe, Camille King, Carol Ryff, Andrew Bersh, Sheher Mohsin, Gayle D. Love, Francesca Nimityongskul, Kristen Malecki, Judith Simcox

**Affiliations:** Department of Biochemistry, University of Wisconsin-Madison, Madison, WI, USA 53706; Integrative Program in Biochemistry, University of Wisconsin-Madison, Madison, WI, USA 53706; Department of Psychology, University of Wisconsin-Madison, Madison, WI, USA 53706; Department of Population Health Sciences, University of Wisconsin-Madison, Madison, WI, USA 53706; Agilent Technologies Inc, Wood Dale, IL, USA 60191

## Abstract

**Rationale:** Cardiovascular disease (CVD) is the leading cause of mortality for women in the USA. Current clinical biomarkers are inadequate to determine CVD risk in women, especially Black women, who disproportionately suffer from CVD.

**Methods:** Clinical data and LC-MS lipidomics from two independent study cohorts were used to identify novel circulating markers of CVD risk in White and Black women. Machine learning assessed predictive efficacy of identified lipids, and targeted oxylipid analysis provided insight into dysregulated inflammatory pathways.

**Results:** Select phospholipids and triglycerides containing acyl chains in the arachidonic acid (ARA) pathway were predictive of systolic blood pressure (BP) after adjusting for biological factors including age, obesity, and glycemic status in White and Black women. Oxylipid levels indicated increased conversion of ARA through the COX and LOX enzymes to pro-inflammatory cytokines in Black women.

**Conclusion:** ARA-containing phospholipid are independent predictors of CVD risk in White and Black women. Predisposition to CVD risk in Black women may further be explained by increased production of pro-inflammatory oxylipids relative to White women, regardless of blood pressure status. Future studies investigating the clinical utility of phospholipid ARA abundance as a marker of CVD risk in White and Black women are warranted.

## Introduction

Cardiovascular disease (CVD) is the leading cause of mortality for women in the USA, accounting for more than 20% of total deaths (1). CVD is most effectively treated through early intervention and diagnosis, which is based on clinical risk factors including plasma cholesterol and triglycerides. Historically, women were excluded from early population studies that identified risk factors of CVD (2). Due to this sampling bias, currently used clinical markers underdiagnose CVD risk in women and precipitate the misconception that CVD is not a major health concern for women (3, 4). Similar risk factors have thus been used to diagnose CVD in women and men despite higher CVD mortality in women since the mid-1980s (5, 6).

In addition to health inequalities due to sex, racial disparities exist for early diagnosis and treatment of CVD. Relative to White women, Black women are more likely to develop CVD earlier in life and are three times more likely to die from hypertension-related diseases (7-9). Despite this disparity in mortality, only 5% of clinical trials for hypertension include Black participants, even though Black people represent 13% of the USA population (10). Consequently, there is poor understanding of the biological factors contributing to CVD in Black relative to White women and a need for studies that focus on early diagnostic markers of CVD in Black women.

Circulating lipids are promising targets for the prediction of CVD. In the clinic, increased plasma low density lipoproteins (LDL) and decreased high-density lipoproteins (HDL) are commonly used to identify CVD risk. However, these markers underdiagnose CVD in women. Pre-menopausal women have higher basal HDL than men of a similar age and Black women with metabolic disease often do not manifest adverse lipoprotein profiles (4, 11). Recent work suggests that focusing on the lipid composition of lipoproteins rather than total LDL and HDL abundance may overcome this discrepancy in CVD diagnosis in women (12). There are sex-specific differences in plasma lipid composition including phosphatidylcholine and phosphatidylethanolamine species, as well as differences associated with race such as higher levels of oxidized fatty acids in Black people (13, 14). Recent work has also linked lipid species to CVD. Use of liquid chromatography-mass spectrometry (LC-MS) based analyses have identified sphingolipids as potential biomarkers of CVD independent of cholesterol, but these studies focused on predominantly male populations (15, 16).The major challenges in applying LC-MS based lipidomics to identify CVD biomarkers rest in the homogeneity of sample participants in population studies and reproducibility between platforms.

We sought to identify novel circulating lipid biomarkers of CVD risk in White and Black women. We analyzed anthropometric data from White and Black women in two independent study cohorts: the Survey of the Health of Wisconsin (SHOW; n=119) and the Midlife in the United States study (MIDUS; n=874). We found that several clinical measurements including circulating cholesterol correlated with high blood pressure (HBP) in White but not Black women. Using untargeted lipidomics and machine learning-assisted predictive modelling, we identified circulating lipids containing arachidonic acid (ARA) as predictive of systolic BP. ARA is an omega-6 fatty acid precursor of bioactive oxygenated lipids (‘oxylipids’), including pro-inflammatory eicosanoids (17, 18). To validate these observations, we performed targeted oxylipid analysis on a subset of MIDUS samples (n=196) and identified oxylipid synthesis pathways that may be differentially active between Black and White women.

Our results indicate that inflammatory pathways related to ARA metabolism are elevated in White and Black women with HBP. Analysis of downstream oxylipids indicates ARA metabolites are higher in Black than White women. These increases in ARA derivatives match with our observations that inflammatory cytokines including c reactive protein (CRP) and interleukin 6 (IL-6) are increased in Black women regardless of BP status. Divergence of inflammatory pathways related to ARA metabolism may affect predisposition to CVD and avenues of treatment for Black women. These findings shed new insight into CVD risk in White and Black women and suggest a need for broader assessment of ARA and inflammatory pathways in the progression of CVD.

## Results

### Baseline differences in clinical parameters of White and Black women

Our goal was to identify novel circulating lipid markers of CVD in White and Black women in the USA. We obtained clinical data on 119 women from SHOW, a human study with banked plasma that represents a diverse intersection of the population including urban and rural, a range of socioeconomic classes, and both Black and White participants (19). SHOW samples were stratified by race and BP status, where systolic > 120 or diastolic > 80 mmHg was considered HBP (**Table 1**) (19, 20). We used HBP as the readout of CVD risk because it is one of the most prevalent markers of CVD risk across multiple populations including White and Black women (21, 22). Median participant age ranged from 50-60 years old and did not significantly differ between White and Black women. Median waist circumference (WC) ranged from 92-103 cm and was above the threshold for abdominal obesity (88 cm) across all groups (23, 24).

**Table 1.** Anthropometric and clinical measurements from the Survey of the Health of Wisconsin (SHOW) indicate metabolic differences between White and Black women. The median [Q1, Q3] of clinical and cytokine data from SHOW participants. ^1^Blood pressure (BP) considered high if systolic/diastolic > 120/80 mmHg. ^2^Kruskal-Wallis rank sum test. For any *P*-values < 0.05, Dunn’s post-hoc test performed and reported with superscript letters after false discovery rate correction. ^3^units of e9 cells/L blood. Abbreviations: BMI, body mass index; WC, waist circumference; BP, blood pressure; HbA1c, hemoglobin A1c; HDL, high density lipoprotein; hsCRP, high sensitivity c reactive protein; IL-6, interleukin 6; sIL-6r, soluble IL-6 receptor; TNF-a, Tumor Necrosis Factor alpha.

| Blood Pressure Status<sup>1</sup> | White Women |  | Black Women |  | *P*-value<sup>2</sup> |
| --- | --- | --- | --- | --- | --- |
|  | Normal | High | Normal | High |  |
| N | 31 | 27 | 25 | 36 |  |
| Age (years) | 50 (38, 58) | 60 (51, 65) | 56 (48, 61) | 54 (49, 60) | 0.069 |
| BMI (kg/m<sup>2</sup>) | 27 (24, 34) | 28 (24, 35) | 29 (26, 35) | 31 (27, 38) | 0.300 |
| WC (cm) | 94 (82, 108) | 92 (83, 116) | 100 (88, 109) | 103 (92, 117) | 0.140 |
| Systolic BP (mmHg) | 109 (102, 114)<sup>b</sup> | 142 (132, 149)<sup>a</sup> | 106 (97, 115)<sup>b</sup> | 140 (124, 147)<sup>a</sup> | **<0.001** |
| Diastolic BP (mmHg) | 68 (62, 72)<sup>b</sup> | 83 (74, 94)<sup>a</sup> | 67 (59, 73)<sup>b</sup> | 84 (80, 89)<sup>a</sup> | **<0.001** |
| White blood cell count<sup>3</sup> | 5.30 (4.10, 6.50) | 5.50 (4.85, 6.35) | 5.60 (4.65, 7.15) | 6.25 (4.75, 8.30) | 0.400 |
| HbA1c (%) | 5.40 (5.30, 5.55) | 5.70 (5.20, 6.10) | 5.80 (5.40, 6.20) | 5.70 (5.38, 6.00) | 0.082 |
| Glucose (mg/dL) | 86 (80, 94)<sup>b</sup> | 93 (88, 97)<sup>a</sup> | 91 (82, 101)<sup>ab</sup> | 94 (86, 99)<sup>a</sup> | **0.040** |
| Total cholesterol (mg/dL) | 191 (172, 212)<sup>b</sup> | 218 (183, 238)<sup>a</sup> | 181 (168, 201)<sup>b</sup> | 184 (160, 225)<sup>b</sup> | **0.012** |
| HDL (mg/dL) | 55 (47, 65) | 54 (43, 68) | 54 (39, 62) | 55 (46, 72) | 0.500 |
| hsCRP (ug/mL) | 5.83 (1.97, 12.2)<sup>b</sup> | 5.64 (1.77, 11.17)<sup>b</sup> | 10.96 (5.93, 16.14)<sup>a</sup> | 14.9 (6.66, 17.94)<sup>a</sup> | **<0.001** |
| Insulin (uIU/mL) | 5 (3, 8)<sup>b</sup> | 6 (4, 9)<sup>ab</sup> | 6 (4, 11)<sup>ab</sup> | 7 (6, 13)<sup>a</sup> | **0.014** |
| IL-6 (pg/mL) | 1.54 (0.83, 3.05) | 1.45 (0.66, 2.76) | 2.15 (0.94, 4.98) | 2.14 (1.57, 3.24) | 0.200 |
| sIL-6r (ng/mL) | 174 (155, 207)<sup>a</sup> | 154 (133, 177)<sup>ab</sup> | 133 (120, 155)<sup>b</sup> | 155 (129, 184)<sup>ab</sup> | **0.003** |
| TNFα (pg/mL) | 0.52 (0.33, 0.89)<sup>a</sup> | 0.35 (0.22, 0.72)<sup>ab</sup> | 0.30 (0.15, 0.53)<sup>ab</sup> | 0.29 (0.13, 0.36)<sup>b</sup> | **0.045** |

Elevated circulating cholesterol is a clinical marker of increased CVD risk (25-27). Consistent with this notion, there was a 14% increase in median total cholesterol in White women with HBP compared to those without (**Table 1**). However, no increase was seen in Black women. Similarly, total cholesterol correlated significantly with systolic BP for White (R=0.35, *P*=0.008), but not Black (R=0.02, *P*=0.852) women. This correlation was driven by non-HDL cholesterol since HDL was unchanged between BP groups (**Fig 1A; Table 1**). These data indicated that circulating cholesterol did not correlate with HBP in Black women.

**Figure 1:**
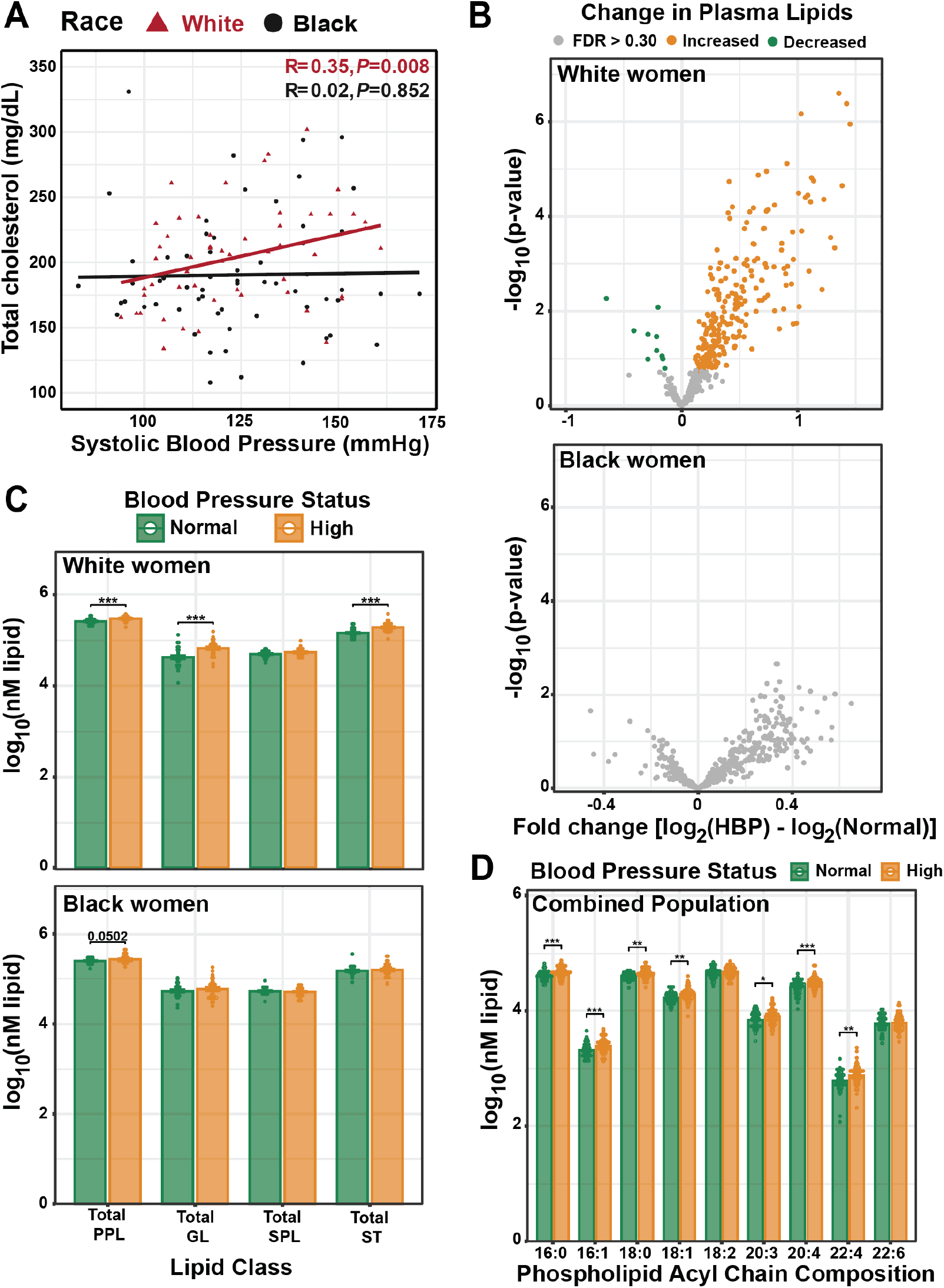
Changes in circulating lipids in women with high blood pressure are dependent on race in SHOW. A) Spearman correlation comparing systolic blood pressure and total cholesterol in White and Black women. B) Volcano plot of plasma lipid changes with high blood pressure (systolic > 120 or diastolic > 80 mmHg) in White and Black women. Change considered significant if FDR ≤ 0.30. C)Total lipid class changes of White and Black women with normal and high blood pressure. D) Abundance of acyl chains in total phospholipids of women with high blood pressure. \**P*<0.05, \*\**P*<0.01, \*\*\**P*<0.001 from pairwise comparison using Welch t-test. Abbreviations: HBP, high blood pressure; PPL, phospholipids; GL, glycerolipids; SPL, sphingolipids; ST, sterol.

To determine if there were differences in basal inflammation due to BP status, we obtained unthawed plasma from SHOW and measured circulating cytokines. Pro-inflammatory CRP was higher (*P*<0.001) in Black women regardless of BP status by approximately 2-fold compared to White women (median>10 ug/mL versus <6 ug/mL; **Table 1**). CRP did not significantly increase due to BP in either race. Similarly, the inflammatory IL-6 cytokine trended higher in Black women but was not significantly different between groups (*P*=0.200; **Table 1**). These data indicated that Black women have increased basal inflammation relative to White women regardless of BP status.

### An acyl chain signature of high blood pressure is present in circulating lipids

Since circulating cholesterol did not correlate with HBP in Black women, we next asked if there were alternative lipid biomarkers of HBP and CVD risk. We performed untargeted lipidomics on plasma from SHOW participants using LC-MS and identified 398 circulating lipid species. There were 239 lipids significantly increased with HBP in White women and the lipid species were quite diverse (**Fig 1B; STable 1**). Strikingly, there were no individual lipid species significantly changed in Black women due to BP status.

Lipid species are categorized by chemical structure and shared synthesis pathways. We classified individual lipids as phospholipids (PPLs), non-PPL glycerolipids (GLs), sphingolipids (SPLs) or sterols (STs). Total PPL, GL, and ST were significantly increased (*P*<0.001) in White women with HBP (**Fig 1C**). Total GL and ST were not significantly increased with BP in Black women, while total PPL was higher (*P*=0.050), matching the trend in White women. These results reflect LDL and HDL levels because the main lipid types in the GL and ST classes triglycerides (TAGs) and cholesteryl esters (CEs) respectively, are major components of circulating cholesterol lipoproteins (28).

Since PPL were increased with HBP in Black and White women, we analyzed acyl chain composition to determine HBP-associated changes. Phospholipids are defined by a glycerol backbone containing a phosphoryl head group and 1-2 acyl chains of varying carbon length and desaturation. Because there were no single phospholipid species changed with BP status in Black women (**Fig 1B**), we asked if there were differences in the acyl chains present in PPLs due to HBP. We observed that many acyl chains were enriched in the PPL pool of women with HBP regardless of race including 16:0, 16:1, 18:0, 18:1, 20:3, 20:4, and 22:4 species (**Fig 1D; SFig 1A**). These trends persisted after stratification by race and were also present in the GL pool (**SFig 1B-C**). Interestingly, several of these lipids had a shared synthesis pathway; both 20:4 and 22:4 are derivatives of the omega-6, essential fatty acid linoleic acid (LA, 18:2), and their synthesis is mediated by a series of desaturases (FADS1 and FADS2) and elongases (ELOVL2/5) (**SFig 1D**). Together, these results indicated the existence of a PPL acyl chain signature of HBP in both White and Black women.

### Traditional clinical measurements correlate with White but not Black women in the MIDUS study

Our SHOW cohort observations indicated that circulating cholesterol correlated with systolic BP in White but not Black women of similar age and obesity status. By profiling plasma lipids, we confirmed that lipid species contained within lipoproteins (i.e. triglyceride and cholesteryl ester species) were not increased in Black women with HBP. However, we were able to identify acyl chain signatures that might indicate CVD risk in White and Black women from SHOW.

To determine if these findings could be extended to a broader population, we analyzed publicly available data from the independent Midlife in the United States (MIDUS) study (**Table 2**) (29). MIDUS is a 30-year longitudinal study aimed at modeling aging US adults and has collected a cache of physiological measures including bone density, fat mass, and inflammatory cytokines. Clinical and lipidomic data was available for 695 White and 179 Black women of which 411 and 137 had HBP, respectively. White women with HBP were significantly older than all other groups (*P*<0.001), reflecting the national trend of higher CVD rates in Black women at younger ages than other races (30). Similar to SHOW, all groups had median WC indicative of obesity, though Black women had notably higher WC (median>98 cm) than White women (<94 cm) regardless of BP status. Total cholesterol was not changed with BP status in Black women but was increased in White women with HBP (**Table 2**). The pro-inflammatory cytokines CRP, IL-6 and tumor necrosis factor alpha (TNF-α) were all significantly (*P*<0.001) increased in White women with HBP compared to normal, but this trend was absent in Black women (**Fig 2A**). However, both CRP and IL-6 were higher in Black women, reflecting a generally elevated inflammation status.

**Table 2.** Anthropometric measurements of women enrolled in the Midlife in the United States (MIDUS) study. The median (Q1, Q3) of clinical data from MIDUS participants.^1^Blood pressure (BP) considered high if systolic/diastolic > 120/80 mmHg. ^2^Kruskal-Wallis rank sum test. Dunn’s post-hoc test performed and reported with superscript letters after false discovery rate correction. Abbreviations: BMI, body mass index; WC, waist circumference; WHR, waist-to-hip ratio; BP, blood pressure; HbA1c, hemoglobin A1c; HOMA-IR, Homeostatic Model Assessment for Insulin Resistance; HDL, high density lipoprotein; LDL, low density lipoprotein.

| Blood Pressure Status<sup>1</sup> | White Women |  | Black Women |  | *P*-value<sup>2</sup> |
| --- | --- | --- | --- | --- | --- |
|  | Normal | High | Normal | High |  |
| N | 284 | 411 | 42 | 137 |  |
| Age (years) | 51 (43, 60)<sup>b</sup> | 60 (52, 68)<sup>a</sup> | 52 (43, 59)<sup>b</sup> | 54 (45, 61)<sup>b</sup> | **<0.001** |
| BMI (kg/m<sup>2</sup>) | 25 (23, 30)<sup>c</sup> | 28 (25, 34)<sup>b</sup> | 32 (28, 39)<sup>a</sup> | 33 (29, 39)<sup>a</sup> | **<0.001** |
| WC (cm) | 84 (74, 94)<sup>c</sup> | 93 (82, 104)<sup>b</sup> | 102 (89, 114)<sup>a</sup> | 99 (90, 112)<sup>a</sup> | **<0.001** |
| WHR | 0.80 (0.76, 0.86)<sup>c</sup> | 0.84 (0.79, 0.89)<sup>b</sup> | 0.88 (0.80, 0.93)<sup>ab</sup> | 0.87 (0.81, 0.93)<sup>a</sup> | **<0.001** |
| Systolic BP (mmHg) | 111 (105, 115)<sup>b</sup> | 136 (127, 146)<sup>a</sup> | 112 (104, 117)<sup>b</sup> | 135 (127, 148)<sup>a</sup> | **<0.001** |
| Diastolic BP (mmHg) | 68 (62, 73)<sup>c</sup> | 76 (71, 83)<sup>b</sup> | 68 (64, 74)<sup>c</sup> | 79 (72, 86)<sup>a</sup> | **<0.001** |
| HbA1c (%) | 5.60 (5.30, 5.90)<sup>c</sup> | 5.80 (5.50, 6.10)<sup>b</sup> | 6.00 (5.43, 6.50)<sup>ab</sup> | 6.10 (5.70, 6.60)<sup>a</sup> | **<0.001** |
| Insulin (uIU/mL) | 8 (5, 13)<sup>c</sup> | 10 (7, 16)<sup>b</sup> | 16 (8, 22)<sup>a</sup> | 14 (8, 22)<sup>a</sup> | **<0.001** |
| Glucose (mg/dL) | 92 (86, 98)<sup>c</sup> | 96 (89, 104)<sup>b</sup> | 94 (90, 106)<sup>ab</sup> | 99 (91, 115)<sup>a</sup> | **<0.001** |
| HOMA-IR | 1.85 (1.17, 3.11)<sup>c</sup> | 2.35 (1.47, 4.10)<sup>b</sup> | 3.91 (1.86, 6.32)<sup>a</sup> | 3.91 (1.95, 6.42)<sup>a</sup> | **<0.001** |
| Total Cholesterol (mg/dL) | 183 (160, 205)<sup>b</sup> | 192 (168, 216)<sup>a</sup> | 178 (143, 198)<sup>b</sup> | 180 (160, 208)<sup>b</sup> | **<0.001** |
| HDL (mg/dL) | 63 (52, 74)<sup>a</sup> | 60 (49, 74)<sup>ab</sup> | 56 (49, 74)<sup>ab</sup> | 56 (47, 71)<sup>b</sup> | **0.033** |
| LDL (mg/dL) | 96 (79, 122)<sup>b</sup> | 104 (84, 124)<sup>a</sup> | 90 (71, 118)<sup>b</sup> | 100 (76, 122)<sup>ab</sup> | **0.014** |
| Triglycerides (mg/dL) | 86 (63, 116)<sup>bc</sup> | 106 (77, 152)<sup>a</sup> | 74 (52, 100)<sup>c</sup> | 94 (68, 132)<sup>b</sup> | **<0.001** |

**Figure 2:**
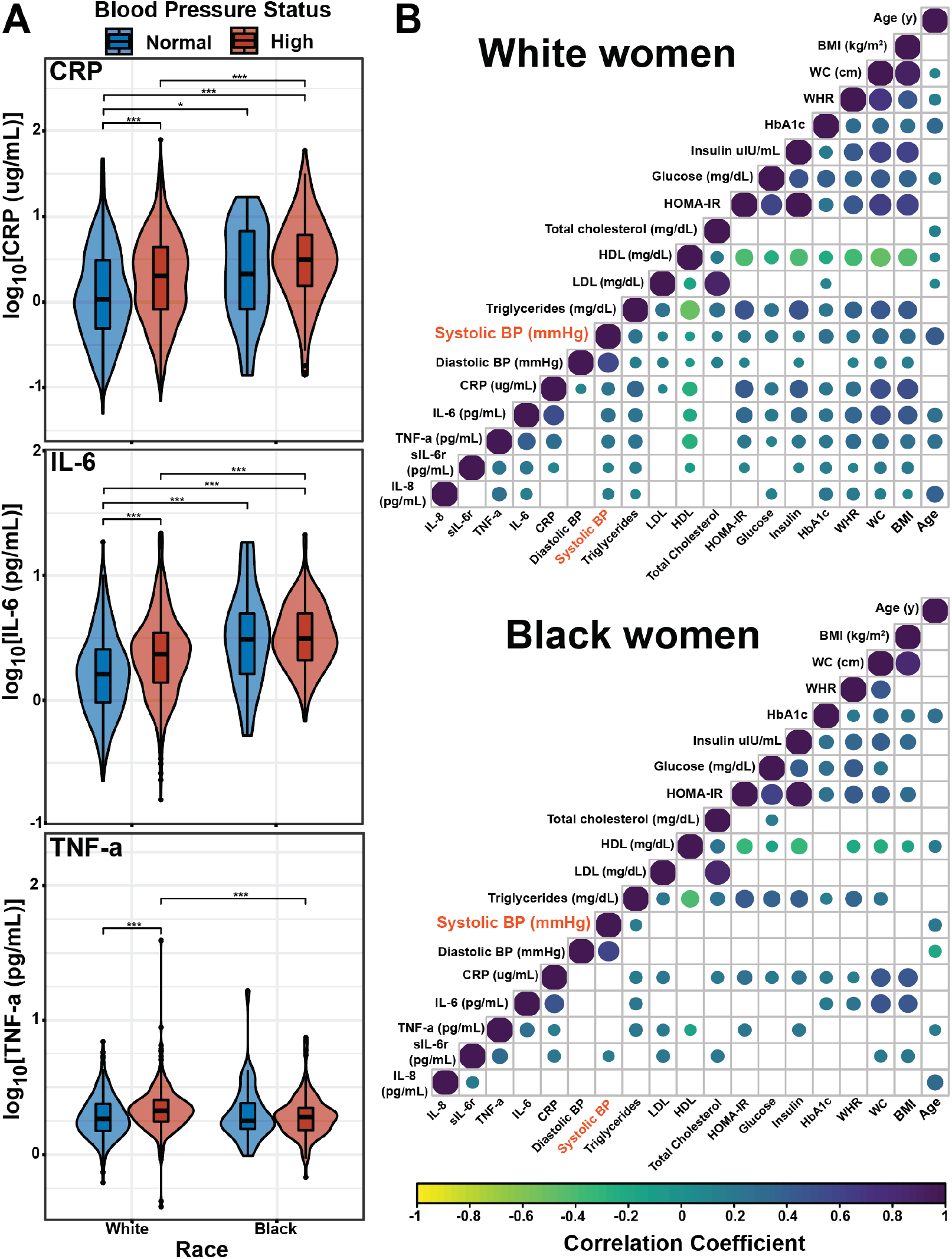
Inflammatory cytokines and traditional clinical markers of metabolic disease correlate poorly with blood pressure in Black women in the MIDUS study. A) Violin plot with boxplot overlay showing changes in distribution and differences of circulating cytokines in White and Black women with normal or high blood pressure. Multiple comparisons considered significant if *P*-values from Dunn’s test < 0.05 after Bonferroni correction. B) Spearman correlations of clinical health and cytokine markers in White and Black women. Only correlations with *P*<0.05 are plotted. \**P*<0.05, \*\**P*<0.01, \*\*\**P*<0.001. Abbreviations: CRP, c reactive protein; IL-6, interleukin-6; TNF-α, tumor necrosis factor alpha; BMI, body mass index; WC, waist circumference; WHR, waist-to-hip ratio; HbA1c, hemoglobin A1c; HOMA-IR, homeostatic model assessment for insulin resistance; HDL, high density lipoprotein; LDL, low density lipoprotein; BP, blood pressure; sIL-6r, soluble IL-6 receptor; IL-8, interleukin-8.

To measure the relationship between clinical measurements, we used Spearman correlations and plotted all significant relationships (*P*<0.05; **Fig 2B**). Many of the results we observed in the White population were expected based on previous literature (4, 27). For example, WC correlated positively with adverse metabolic effects such as increased hemoglobin A1c (HbA1c), glucose, triglycerides, CRP, and IL-6 (*R*=0.30-0.80) and inversely with HDL (*R*=-0.48). Similarly, both systolic and diastolic BP positively correlated with total cholesterol, LDL, and all inflammatory cytokines (*R*=0.10-0.30). In contrast, there was a general lack of significant correlations between variables in the Black population. The clearest example was that neither systolic nor diastolic BP correlated significantly with total cholesterol, LDL, WC, CRP, or IL-6 in Black women (**Fig 2B**). Together, these findings support and expand on our findings from SHOW that 1) basal inflammatory cytokines are increased in Black women regardless of BP status; and 2) traditional clinical markers of CVD risk such as total cholesterol correlate with systolic BP in White but not Black women.

### Arachidonic acid-containing lipids are increased in White and Black women with high blood pressure

We next turned to untargeted lipidomics data from MIDUS to identify signatures of CVD risk applicable to both White and Black women. One of the major challenges in application of broad lipidomics is the disparity in identification and quantification between instrumentation platforms. MIDUS was selected because it was collected using a different analytical platform to ensure observations were translatable across instrumentation (31). Lipidomics on SHOW samples was performed on an Agilent LC/MS, whereas MIDUS data was collected by Metabolon Inc. using the SCIEX Lipidyzer that identified 381 lipid species. Most measured lipid species (280 lipids) were increased with HBP in the White population (**Fig 3A; STable 1**). In agreement with SHOW findings, there were no lipids significantly changed with HBP in the Black population.

**Figure 3:**
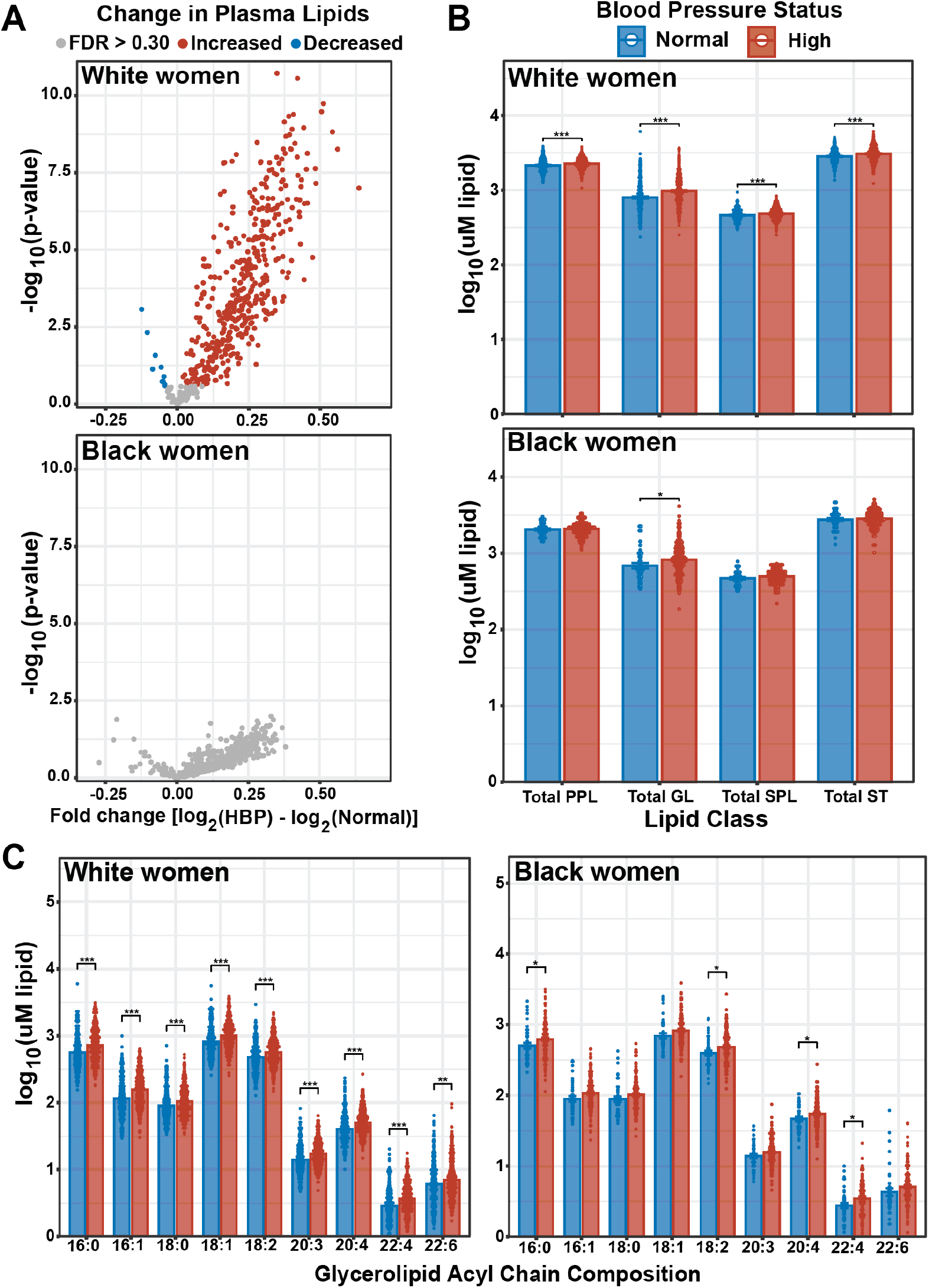
Differences in lipid acyl chain but not lipid species are associated with high blood pressure in Black and White women in MIDUS. A) Volcano plot of lipid changes with HBP (systolic > 120 or diastolic > 80 mmHg) in White and Black women. Change considered significant if FDR ≤ 0.30. B) Comparison of total lipid class changes in plasma lipids of White and Black women with normal or high blood pressure. C) Comparison of the prevalence of the most abundant acyl chains in total glycerolipids of White and Black women with HBP. \**P*<0.05, \*\**P*<0.01, \*\*\**P*<0.001 from pairwise comparison using Welch t-test. Abbreviations: HBP, high blood pressure; PPL, phospholipids; GL, glycerolipids; SPL, sphingolipids; ST, sterol.

When we compared lipid class changes, all classes were significantly increased in the White population with HBP (*P*<0.001), but only total GL were elevated with HBP in Black women (*P*<0.05; **Fig 3B**). These observations in the Black population differed from SHOW, where total PPL but not GL were increased with HBP (**Figure 1C**). Nevertheless, when we compared the GL acyl chain composition for White and Black populations, several acyl chain signatures present in SHOW persisted including elevated 16:0, 18:2, 20:4 and 22:4 in HBP (**Fig 3C**). The 20:4 acyl chain is intriguing since the most prevalent 20:4 fatty acid is the omega-6 ARA. ARA has been extensively studied as a precursor to many downstream metabolites that mediate inflammation including eicosanoids. Additionally, 18:2 and 22:4 exist in the biosynthetic pathway of ARA. 18:2, most often LA, is elongated and further desaturated to form ARA, while 22:4 is an elongation product of ARA and a source of ARA after undergoing one round of beta oxidation (**SFig 1D**).

Further, 22:4 was increased in the GL pool of SHOW samples as well (**SFig 1A & C**). These data indicated that lipids containing ARA and related acyl chains may be indicators of, and potentially mediators of, CVD risk in White and Black women.

### Common lipid species are predictive of systolic blood pressure despite differences in analytical platforms between SHOW and MIDUS

Acyl chains can be incorporated into several lipid classes including PPLs (1-2 acyl chains), GLs (1-3 acyl chains), and CEs (1 acyl chain). Although we observed increased ARA in women with HBP in both the SHOW and MIDUS lipidomics, the lipid classes contributing to increased ARA were different between studies (**Fig 1D & 3C; SFig 1A & C**). We hypothesized that this may reflect differences in the ARA-containing lipids measured by the lipidomic platforms. We compared distribution of ARA abundance across all measured lipids between SHOW and MIDUS and observed that the majority of ARA containing lipids measured in the MIDUS study were from CEs (33% total abundance), phosphatidylcholines (PCs; 51%), and TAGs (8%)(**Fig 4A**). In SHOW, CEs and PCs were also major contributing lipids to the total 20:4 abundance, but the PC contribution was 23% less than in MIDUS. In SHOW, phosphatidylethanolamines (PEs; 14%) contributed the third highest ARA abundance while TAGs were only 2% of the total abundance. This could be explained by differences in the number of ARA containing species detected in each lipid class as twice the number of PEs were detected in SHOW than MIDUS (8 versus 4 lipids) while there were 20 more TAGs measured in MIDUS than SHOW (27 versus 7) (**Fig 4A**). Altogether, more PPLs containing ARA were detected in SHOW whereas more GLs were detected in MIDUS, explaining why ARA enrichment was detected in different lipid classes between the two studies. These findings highlight how analytical differences in lipidomic measurements can affect the detected patterns and relationships with other variables across studies, reflecting the importance for standardization of lipidomics measurements to improve translational application.

**Figure 4:**
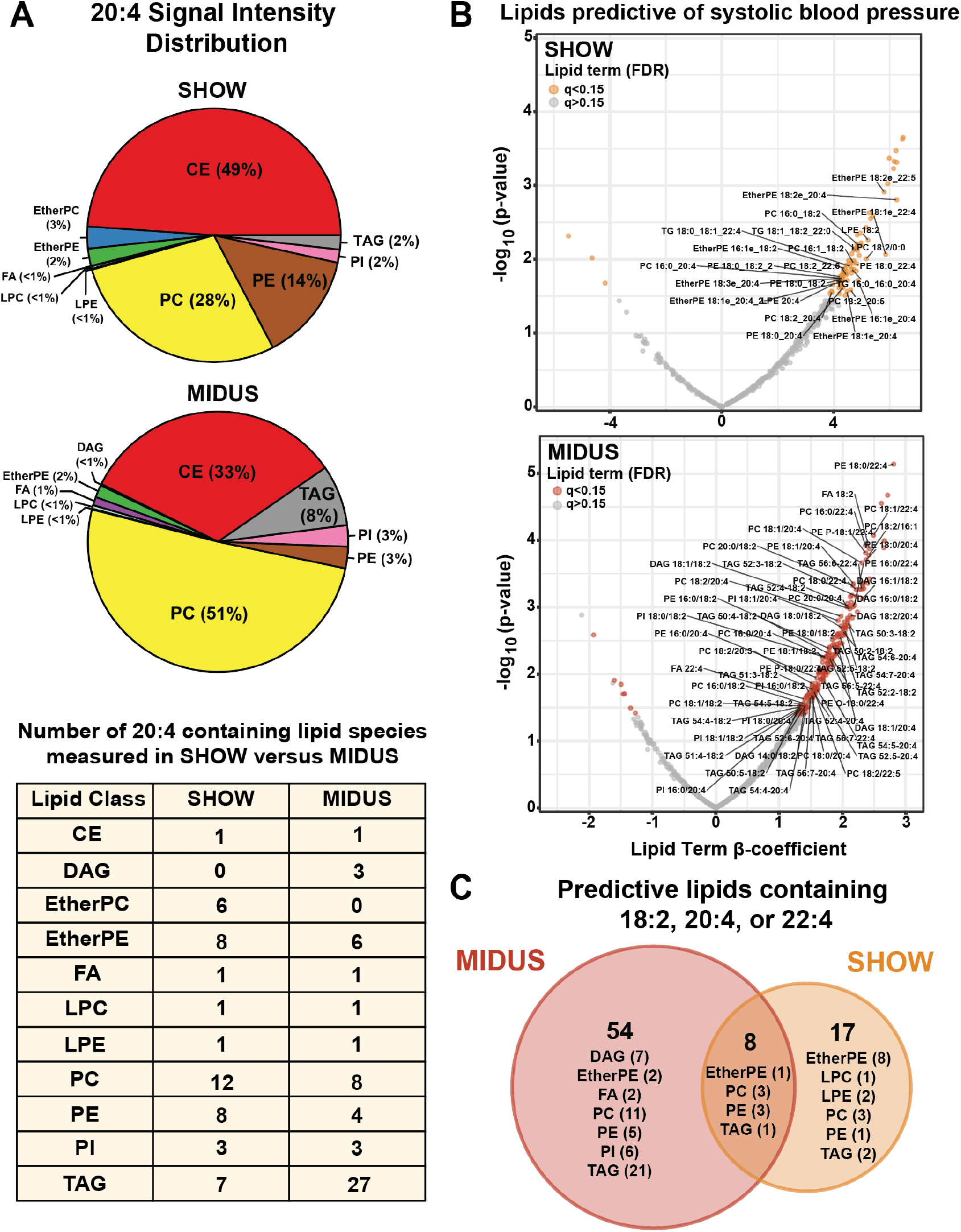
Arachidonic acid-containing lipids detected in both SHOW and MIDUS are predictive of systolic blood pressure after adjusting for covariates. A) Pie chart comparing the average signal distribution of 20:4 detected lipid classes in the entire SHOW and MIDUS populations. The number of individual lipid species constituting each class is provided in the table. B) Multiple regression analyses for each lipid species (beta coefficient and p-value of the lipid term) predicting systolic blood pressure are plotted after adjusting for race, age, waist circumference, and c reactive protein. Each lipid was regressed separately within the respective studies. C) Venn diagram comparing lipid species positively predictive of systolic blood pressure from Figure 4B and containing either 18:2, 20:4, or 22:4 is shown with the lipid classes in each portion and number of lipid species in parentheses. Abbreviations: CE, cholesteryl ester; DAG, diglyceride; FA, fatty acid; PC, phosphatidylcholine; PE, phosphatidylethanolamine; LPC, lysoPC; LPE, lysoPE; PI, phosphatidylinositol; TAG, triglyceride.

Next, we sought to directly assess lipid species predictive of systolic BP. Because biological factors such as age, obesity, glucose homeostasis and inflammation can affect circulating lipids, we regressed each lipid species detected in either SHOW or MIDUS to systolic BP with race, age, WC, HbA1c and CRP as covariates. The β-coefficient and p-values for the lipid terms are reported in **Fig 4B**. In both studies, several lipids in the ARA pathway–those containing 18:2, 20:4 and 22:4 chains– were positively and significantly (*P*<0.05) predictive of systolic BP (**Fig 4B; STable 2**). When we directly compared lipids containing 18:2, 20:4 or 22:4 acyl chains between the two studies, eight species were measured in both studies and predictive of systolic BP (**Fig 4C; STable 2**). These lipids were PC 16:0_18:2, PC 16:0_20:4, PC 18:2_20:4, PE 18:0_18:2, PE 18:0_20:4, PE 18:0_22:4, EtherPE 18:1_22:4 and TAG 52:4 containing one 20:4 chain. Given that many ARA-containing lipids were detected in only one study and these lipids were predictive of systolic BP after accounting for several other biological factors, these findings strongly suggested that ARA and related lipids are candidate markers of CVD risk in White and Black women.

### Machine learning identifies 18:2, 20:4 and 22:4 lipids as predictive of systolic blood pressure

We decided to focus on the MIDUS study for follow-up analyses due to a larger sample size of both Black and White women. Multiple linear regressions had identified 62 lipids in the ARA pathway as predictive of systolic BP including those containing 18:2, 20:4 or 22:4 acyl chains. When we performed a regression using these ARA pathway lipids to predict systolic BP, the model was highly significant (*P*<0.001) with an R^2^=0.25 (**SFig 2A**). Adding back covariates (age, WC, HbA1c and CRP) improved the regression with a new R^2^=0.33 (**SFig 2B**). These results indicated that 18:2, 20:4 and 22:4 lipids could predict systolic BP independent of other clinical markers including age and WC, and that this predictive value was particularly robust for Black women.

We next used an orthogonal method to predict systolic BP with the additional goal of limiting the number of independent variables to the fewest possible lipids. To accomplish this, we turned to machine learning and used the lasso regression method, which has been successfully employed as a feature selection technique for lipidomic data (16). After training our model on 70% of the MIDUS data, we successfully predicted systolic BP of the remaining 30% of the data with similar performance as the training data (**Fig 5A**). The overall model mean average error (MAE) was 12.97 for the training dataset and 13.37 for the test data, with overall R^2^ values of 0.29 and 0.22, respectively. As previously observed (**SFig 1A-B**), the model was more accurate in predicting BP for White than Black women.

**Figure 5:**
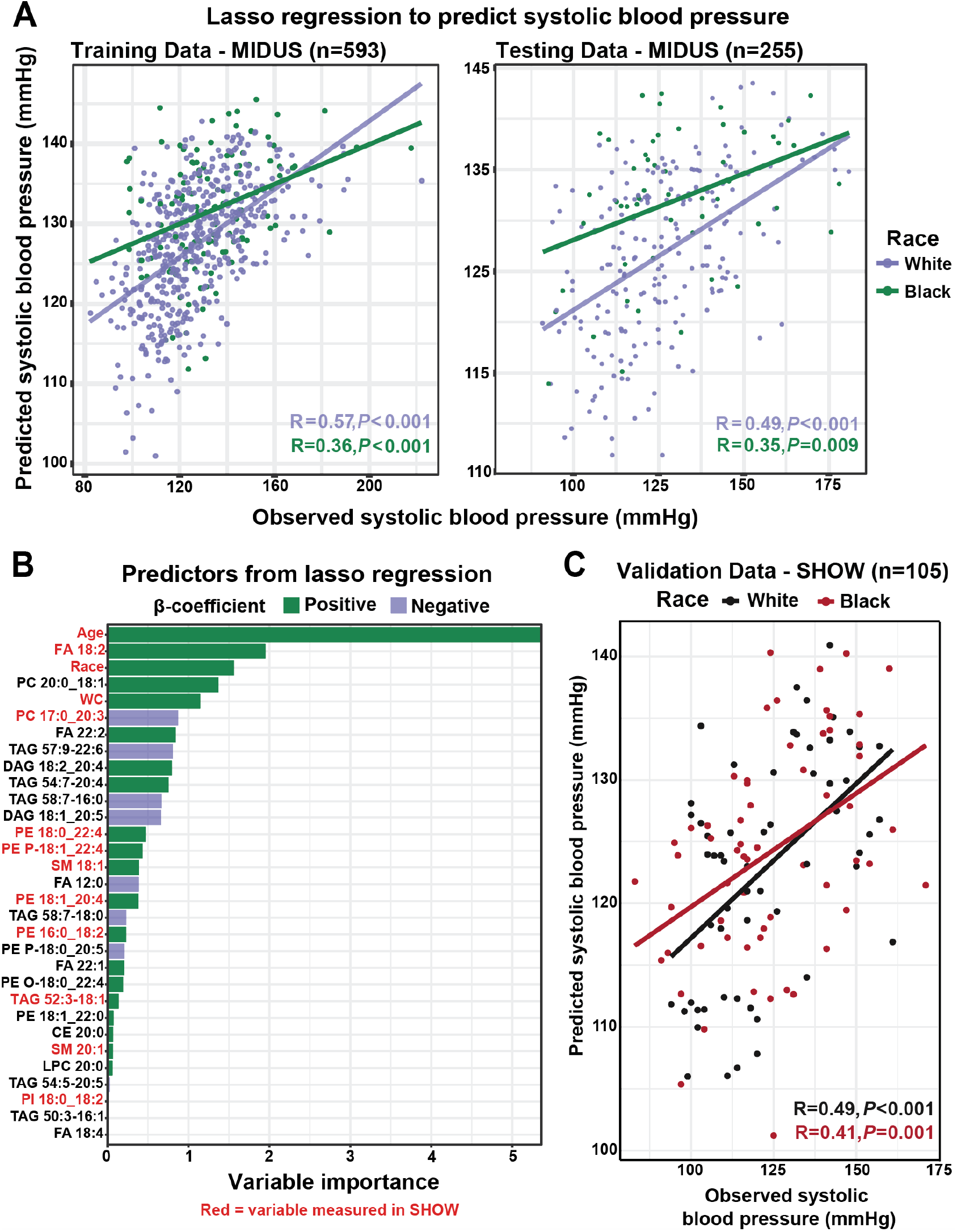
Machine learning identifies twenty-eight lipids significantly predictive of systolic blood pressure. A) Scatterplot of systolic blood pressure predictions using the final lasso regression is plotted against the actual blood pressure for the MIDUS training and test data sets. The training was used to optimize the model whereas test data was sequestered throughout optimization. B) Variable importance plot of the final lasso model predictors is presented. Variables in red were measured in the SHOW cohort and used for validation of the final model. C) Scatterplot of systolic blood pressure predictions using results from the final lasso regression is plotted against the actual blood pressure for the entire SHOW dataset as independent cohort validation of findings. Note that only variables in red from Figure 5B could be used because the other variables were not measured in SHOW. Abbreviations: FA, fatty acid; PC, phosphatidylcholine; WC, waist circumference; TAG, triglyceride; PE, phosphatidylethanolamine; PE P- or PE O-, ether PE; SM, sphingomyelin; CE, cholesteryl ester; LPC, lyso PC; PI, phosphatidylinositol.

The final lasso regression consisted of 31 variables (**Fig 5B**). The most significant predictor was age, while circulating 18:2 was the most important lipid predictor. Interestingly, CRP and HbA1c were not part of the final model indicating basal inflammation and glucose homeostasis may not be major predictors of BP in women. Approximately one third of the final lipid predictors contained either 18:2, 20:4, or 22:4 and all of them were positive predictors of systolic BP. Though most lipid variables in the final lasso regression were not measured in the SHOW dataset, we validated the model with the subset that were present in SHOW. Since SHOW and MIDUS are independent cohorts with different analytical techniques used for lipidomics measurements, we reasoned that similar trends would increase confidence in our findings. Using the thirteen lasso predictors measured in SHOW (6 containing 18:2, 20:4 or 22:4), we had strong predictability of systolic BP in both Black and White women (MAE=13.86, R^2^=0.215; **Fig 5C**).

Together, our results indicated that ARA pathway lipids predicted systolic BP independent of age, race, WC, diabetes status and basal inflammation. Finally, we sought to understand how these lipids may function to increase CVD risk with a focus on downstream oxylipids and regulation of inflammatory pathways.

### COX and LOX-derived oxylipids are associated with systolic blood pressure

To gain insight into ARA pathway derivatives that mediate inflammation and underlie CVD risk, we obtained 196 unthawed plasma samples from MIDUS and analyzed them for oxylipids using a targeted LC-MS method. The samples were from 96 White women (n=51 with HBP) and 100 Black women (n=60 with HBP). Oxylipids are signaling lipids that are produced from polyunsaturated fatty acids and regulate inflammation (18). Oxylipids derived from the ARA pathway include prostaglandins, leukotrienes, and hydroxyeicosatetraenoic acids (HETEs) (32). Our method screened for 168 oxylipids, 89 of which met our inclusion criteria and were used for further analysis (see Methods; **STable 3-4**).

The majority of oxylipids analyzed originated from LA, ARA, eicosapentaenoic acid (EPA), or docosahexaenoic acid (DHA) and were produced through shared enzymatic pathways (**STable 3**) (33, 34). We correlated each oxylipid with systolic BP in White and Black populations to identify underlying relationships between BP and oxylipids of shared synthetic pathways (**Fig 6A**). The majority of oxylipids significantly correlated with systolic BP in the White population were derived from ARA. Most of these oxylipids were associated with COX enzyme metabolism, with the exception of 14,15-DiHETE, 12-oxoETE and 12-HETE. Unexpectedly, all correlations between systolic BP and oxylipid levels were negative associations. This sharply contrasted with the expected positive association indicative of the traditional pro-inflammatory states typical of CVD.

**Figure 6:**
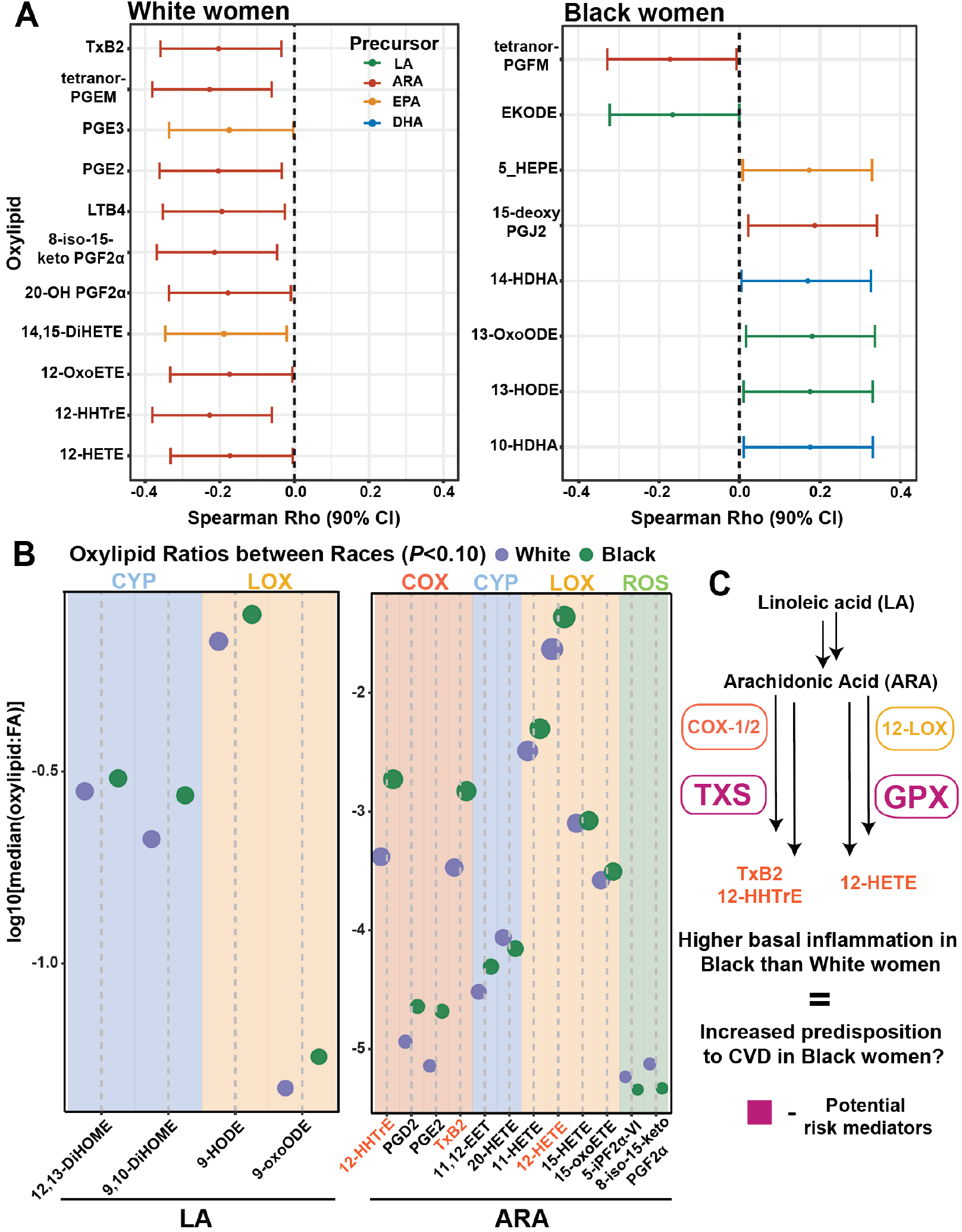
Arachidonic acid metabolism to oxylipids diverges between White and Black women. A) Spearman correlations between systolic blood pressure and oxylipids are given for White and Black women from MIDUS. Precursor fatty acid is indicated by line color. 90% confidence interval was used. B) The ratio of individual oxylipid to precursor fatty acid was taken and compared between White and Black women. Median linoleic and arachidonic acid-related ratios significantly different (*P*<0.10) between race are plotted and the major enzymatic branch indicated. C) Pathways of interest, based on largest median difference and most abundant oxylipid from B, are detailed. Enzymes in the pathway are boxed. Abbreviations: LA, linoleic acid; ARA, arachidonic acid; EPA, eicosapentaenoic acid; DHA, docosahexaenoic acid; CYP, cytochrome P450; LOX, lipoxygenase; COX, cyclooxygenase; ROS, reactive oxygen species; FA, fatty acid; TxB2, thromboxane B2; 12-HHTrE, 12-hydroxyheptadecatrienoic acid; 12-HETE, 12-hydroxyeicosatetraenoic acid; TXS, thromboxane synthase; GPX, glutathione peroxidase. Oxylipid abbreviations are provided STable 3.

In Black participants, there were eight significant oxylipid correlations with systolic BP. Unlike the White population, there was no specific fatty acid signature of correlated oxylipids and most of the metabolites originated from lipoxygenase (LOX), not COX, metabolism. In agreement with previous research, these results indicated that the relationship between oxylipids and CVD was related to broader enzymatic pathways, not individual oxylipid species (34). Interestingly, there seemed to be a difference in the manifestation of these relationships based on race. In White women, systolic BP was inversely related to COX-derived ARA oxylipids, whereas LOX-derived metabolites from multiple fatty acids positively correlated with systolic BP in Black women.

Direct comparison of oxylipid levels of White women with and without HBP identified two compounds significantly decreased (*P*<0.05) with HBP – 8-iso-15-keto PGF2α and 20-OH PGF2α - and none increased (**STable 3**). After FDR correction, no oxylipids reached significance (q<0.30; **SFig 3A**). There were no oxylipids reaching either significance threshold when comparing BP groups in the Black population (**SFig 3B; STable 3**). These data indicated that there were no overt differences in individual, free oxylipids between women with or without HBP irrespective of race. During conditions such as CVD in which individuals are at generally higher inflammation states, changes in flux through oxylipid metabolic pathways may be spread out across many analytes rather than individual ones. Therefore, we investigated whether differences in broad oxylipid synthesis pathways may underlie BP-associated changes.

### Race-based differences in circulating oxylipids indicate different pathways mediate CVD risk between White and Black women

We had identified several oxylipids associated with systolic BP, however, these relationships diverged for White and Black women based on both oxylipid fatty acid precursor and major enzymatic pathway. This led us to focus on identifying specific enzymes underlying differences in CVD risk in Black women using oxylipid levels. Identification of these targets would provide novel biological insight into disease pathology and treatment options in these populations, especially since enzyme activity can be altered by drugs, diet, and other environmental factors (35, 36).

We first compared changes in oxylipid abundance in White versus Black women regardless of BP status (**SFig 3C**; **STable 3**). There were 43 oxylipids significantly different between races after FDR adjustment. The majority of these oxylipids (35) were higher in the White population. Seeking insight into enzymatic pathways mediating these differences, we took the ratio of free oxylipids (product) to the fatty acid of origin (precursor) and compared the median of all ratios significantly higher (*P*<0.10) in a particular race (**Fig 6B; SFig 3D**). The majority of LA and ARA-derived oxylipid ratios were higher in the Black population. The greatest ratio fold changes were the COX-derived 12-HHTrE- and thromboxane B2 (TxB2)-to-ARA ratios. Both of these oxylipids are derived from the sequential action of COX-1/2 and thromboxane synthase (TXS) followed by non-enzymatic processing (**Fig 6C**). The 12-LOX derived 12-HETE-to-ARA ratio was also significantly higher in Black than White women. 12-HETE was one of the most abundant oxylipids measured in our analysis (median plasma concentration=12.5 ng/mL versus 0.381 for TxB2) and may be a determinant in the inflammatory differences between Black and White women (**Fig 6C & 2A; Table 1**). Taken together, these data indicated that the COX-1/2-thromboxane synthase and 12-LOX-glutathione peroxidase (GPX) pathways may be differentially active between races and explain some of the differences in basal inflammation - and predisposition to CVD - between Black and White women (**Fig 6C**).

## Discussion

CVD is the leading cause of death for women in the USA. The majority of clinical diagnostics used to assess CVD risk were developed from population studies predominantly composed of White men and have been shown to underdiagnose CVD in women (2). We sought to identify markers of CVD risk targeted towards White and Black women, as Black women suffer disproportionately from CVD (8). Through biochemical analysis of two independent population cohorts, SHOW and MIDUS, we confirmed that traditional clinical markers of CVD were poorly correlated with systolic BP (**Fig 1A & 2B**). Untargeted lipidomics analysis of each cohort, using separate analytical platforms, revealed that lipids containing ARA predicted systolic BP after accounting for other biological factors including age and obesity (**Fig 4B & 5**). After performing targeted oxylipid analysis on a subset of MIDUS samples, we found differential ARA metabolism in White and Black women that may underlie predisposition to CVD risk (**Fig 6**). These findings provide novel insight for CVD risk in Black women and illuminated specific pathways for further study to understand disease progression, prevention, and treatment.

Important aspects of our experimental design were the selective inclusion of White and Black women and agreement of our findings in two independent study cohorts. This allowed us to identify population specific markers of cardiovascular health both unique to and shared between Black and White women. We identified lipids containing ARA, particularly PC and PE species, as predictive of systolic BP in White and Black women after adjusting for age and several other factors (**Fig 4B-C; Fig 5B; STable 2**). Biologically, PC and PE lipids are known targets of circulating phospholipases which release ARA for downstream signaling to mediate inflammation during CVD (37). Our regressions further indicated that ARA-containing lipids predict systolic BP in women independent of covariate adjustment, particularly for Black women (**SFig 2**). This latter finding is in agreement with previous observations that the fatty acid desaturase allele most prevalent in the Black population increases production of ARA (38). Another potential driver of increased ARA levels could be from the diet, however, dietary consumption of LA poorly correlates with ARA levels in the serum (39, 40). Remarkably, the relationship between ARA lipids and BP was detected in both study cohorts despite the use of lipidomic methods favoring the identification of different lipid species within each analysis (TAGs in MIDUS and PPLs in SHOW; **Fig 4A**). Taken together, our results form a strong basis for further exploration of ARA lipids as CVD risk factors in women.

ARA metabolism is a key mediator of inflammation. An important marker of systemic inflammation is circulating CRP, which is higher in women than men regardless of ethnicity (41). We observed that CRP is also higher in Black women compared to White women, which aligns with previously published studies (**Table 1; Fig 2A**) (42). Our data further suggests that CRP does not increase with HBP in Black women and is not predictive of systolic BP in women (**Fig 2A-B; Fig 5B**). Beyond the lack of correlation with HBP in Black women, CRP increases are not unique to CVD and are seen in a number of metabolic disease including type 2 diabetes (43).

We observed that the ARA derived pro-inflammatory 12-HETE and TxB2 oxylipids are elevated in Black women irrespective of BP status. Previous work has identified elevated 12-HETE as a mediator of hypertension in other populations and it has been suggested that downstream effects of 12-HETE may induce thromboxane A2 (TxA2) production to increase CVD risk (44, 45). TxA2 is an active vasoconstrictor that is metabolized to TxB2 and eventually 12-HHTrE. Low-dose aspirin use –which lowers TxA2 and therefore TxB2 through COX-1 inhibition– is associated with lower CVD risk in White but not Black women, though aspirin use has mild effects in reducing CVD in the population at large (46, 47). Hence, it remains unclear if and how COX-1/2 inhibition influences CVD risk, though our data suggests that this pathway requires further study for disease prevention in Black women.

Our study has several limitations due to its cross-sectional nature and pertaining to data availability that should be the basis of future study designs. We used HBP as an indirect readout of CVD risk. The benefits of HBP use include its high association with CVD in Black and White populations and measurement of BP as a standard of care for primary care physicians (21, 48). However, a direct measure of CVD outcomes will be necessary in future validation studies. We were also unable to account for medication use due to low response rates in the SHOW and MIDUS questionnaires. Drugs such as NSAIDs impact pathways related to eicosanoid generation, which are a type of oxylipid, and this may have affected our findings. In addition, we could not account for the use of drugs to treat CVD such as statins which likely influenced our ability to detect certain lipids and clinical measurement relationships. It is worth noting that hypertension rates in Black Americans are higher than White Americans independent of therapeutic treatments (48).

The low survey response rates also impacted our ability to account for menopausal status. It has been observed that post-menopausal women are at higher CVD risk though it is unclear why (4). It is likely that some of the variation from this effect was accounted for in our regressions through the age covariate. Additionally, we do not have information on diet, the role of which cannot be overstated when considering health outcomes. We identified ARA and related lipids as markers of CVD risk. ARA is an omega-6 fatty acid which is taken solely from the diet both directly and through elongation and desaturation of the essential fatty acid LA (**SFig 1D**). Previous studies have demonstrated that dietary intake of ARA and LA do not correlate with serum ARA levels (43). Additionally, micronutrients are effectors of inflammatory status and oxylipid levels and these have not been quantified in the MIDUS population (49, 50). Our oxylipid data indicates a higher basal inflammation in Black women compared to White, and it is important to note that Se intake is inversely correlated with production of pro-inflammatory oxylipids such as ARA-derived HETEs (50). Future studies should include information on diet to better delineate biological versus environmental factors in mediating CVD risk in women.

In conclusion, we have identified increases in ARA-containing lipids in White and Black women with HBP. We further found these lipids to be predictive of systolic BP in two independent study cohorts. Analysis of oxylipids downstream of ARA indicates differential production of pro-inflammatory oxylipids in Black and White women, providing insight into novel markers and mechanisms mediating CVD in these understudied populations.

## Data Availability

All code used in this analysis will be made freely available on Github (RJain52). Due to IRB restrictions, all data collected for the MIDUS study must be requested through the appropriate process (http://midus.wisc.edu) while SHOW data is available through SHOW study (show.wisc.edu).

http://show.wisc.edu

## Author Contributions

RJ and JS designed the project. RJ, JD, SM, and PG performed the experiments, and RJ, JD, CK, and JS analyzed the data. CR, AB, CC, and KM collected the human samples and maintained the biobank. RJ, JD and JS prepared the manuscript.

## Acknowledgments

We would like to thank the Mass Spectrometry Core at the UW-Madison Biotechnology facility for instrument use for untargeted lipidomics of the SHOW samples. We would also like to thank Dudley Lamming and members of the Simcox Lab including Gina Wade and Isabella James for providing feedback on the manuscript.

## Data Availability

All code used in this analysis will be made freely available on Github (RJain52). Due to IRB restrictions, all data collected for the MIDUS study must be requested through the appropriate process (http://midus.wisc.edu).

## Methods

### Reagents and standards

The following reagents were purchased for lipid analysis and organic solvents were high performance liquid chromatography (HPLC) grade or higher: Dulbecco’s phosphate buffered saline (DPBS; ThermoFisher Scientific, Waltham, MA, USA #1490235), methanol (MeOH;), methyl tert-butyl ether (MTBE; Fisher Scientific, ThermoFisher #E127-4), isopropanol (IPA; Fisher #A461-4), acetonitrile (MeCN; Honeywell, Charlotte, NC, USA #LC015-4), water (Honeywell #LC365-4), hexane (Millipore Sigma, Burlington, MA, USA #1037012500), methyl formate (Sigma-Aldrich, St. Louis, MO, USA #291056-1L), formic acid (Fisher #UN1779), and acetic acid (Fisher #A113-50). Butylated hydroxy-toluene (BHT) was purchased from Sigma-Aldrich (#B1378-500G) and ammonium formate from Fisher (#A115-50).

SPLASH I LipidoMix internal standard for untargeted lipidomics was purchased Avanti (Alabaster, AL, USA #330707). For oxylipid analysis, the follow standards were purchased from Cayman Chemical (Ann Arbor, MI, USA): DHA oxylipin mix (#22280), lipoxin mix (#19412), SPM D-series mix (#18702), linoleic acid mix (#20794), leukotriene B4 mix (#22640), primary COX and LOX mix (#19101), secondary prostaglandins mix (#19422), EPA CYP450 mix (#21394), oxidized lipid HPLC mix (#34004), 14(15)-EET (#10007263), maresin-1_d5_ (#26969), 12(13)-diHOME_d4_ (#10009994), DHA_d5_ (#27357), EPA_d5_ (#27358), AA_d11_ (#24919), 14(15)-diHET_d11_ (#0528483), lipoxin A4_d5_ (#24936), 20-HETE_d6_ (#24923), prostaglandin E2_d4_ (#10007273), prostaglandin D2_d4_ (#10007272), 9-HODE_d4_ (#25368), leukotriene B4_d4_ (#29629), thromboxane B2_d9_ (#23579), and 14(15)-EET_d11_ (#26970).

### MIDUS sample data

Publicly available data from MIDUS was obtained from http://midus.wisc.edu. All specimen collection and testing for MIDUS are approved by the Health Sciences Institutional Review Board at the University of Wisconsin-Madison as well as the Institutional Review Boards at the University of California-Los Angeles and Georgetown University. This includes clinical and cytokine measurements. Lipidomic analysis was performed by Metabolon as previously described (51, 52). We would like to note that for TAGs in particular, the Metabolon platform reports the acyl chain distribution of fatty acids in the TAG pool but not individual species because the overall acyl chain composition of TAGs is not reported (e.g. TAG 56:4-20:4 is reported rather than the individual lipid species such as TAG 18:0_18:0_20:4). This is also true for ceramide species in which the sphingoid bases are not reported. This annotation format leads to duplications in identification and difficulty with summative assessment of total species contribution. These factors were taken into consideration during subsequent analysis. Native Metabolon reporting (e.g. TAG 56:4-20:4) was treated as a sum value (‘total 20:4 in TAGs with the 56:4 sum composition’) and an ‘individual lipid value’ was calculated where TAG 56:4 was calculated by adding all 56:4-X (X= acyl chain) lipids and dividing by three, the total number of acyl chains in a TAG.

For targeted oxylipid measurements, 196 plasma samples, never thawed, were obtained from MIDUS. Details on oxylipid protocols are described below. All sample collection occurred prior to the COVID-19 pandemic and samples were stored at -80°C prior to analysis. Downstream processing and handling of data is further described in the *Data analysis* section.

### Survey of the Health of Wisconsin (SHOW) sample data

Independent validation of MIDUS findings were conducted using 119 plasma samples from women enrolled in the SHOW study (show.wisc.edu; IRB #2013-0251, #2020-0752). Samples were selected to include a similar number of women with high and normal blood pressure within the respective races. Cytokine and untargeted lipidomics data were generated for this study as described below. All samples were collected prior to the COVID-19 pandemic and samples were stored at -80°C until analyses.

### ELISA measurements of cytokines

Commercially available ELISA kits were used for all measurements from the SHOW samples. The ELISAs were purchased from ThermoFisher Scientific: C-reactive protein (CRP; Invitrogen #KHA0031), insulin (Invitrogen #KAQ1251), tumor necrosis factor α (TNF-α; Invitrogen #BMS223HS), interleukin-6 (IL-6; Invitrogen #BMS213HS), soluble IL-6 (sIL-6; Invitrogen #BMS214INST), IL-8 (Invitrogen #KHC0081), and IL-10 (Invitrogen #BMS215HS). Manufacturer instructions were followed.

### Untargeted lipidomics extraction

Plasma samples from SHOW were extracted for untargeted lipidomics using the method of Matyash *et al*. All work was done on ice and centrifugation was at 4°C. To ceramic bead tubes (Qiagen, Hilden, Germany #13113-50), 250 μL DPBS, 215 μL MeOH containing 10 μL SPLASH internal standard and 40 μL plasma was added and mixed by inversion (53). 750 μL MTBE was added and samples were homogenized 2×30 s using the TissueLyzer II (Qiagen #85300). Samples were incubated on ice for 15 min then centrifuged at 16,100xg for 5 min. The top organic layer was transferred to a microfuge tube and dried under a gentle stream of nitrogen at room temperature. Samples were reconstituted in 150 μL of IPA and stored at -80°C until analysis.

### LC-MS untargeted lipidomic analysis

LC-MS was performed for untargeted lipidomics of SHOW samples as previously described, with minor modifications (54). An Agilent (Santa Clara, CA, USA) 1290 Infinity II LC coupled to an Agilent 6546 quadrupole time-of-flight MS was used for analysis. The LC was equipped with an Agilent InfinityLab Poroshell 120 C18 column (3.0×50 mm, 2.7 μm) operated at 50°C with a constant flow rate of 0.500 mL/min and the following gradient: start at 15% B to 30% B at 2.4 min, to 48% B at 3 min, to 82% at 13.2 min, to 99% at 13.8 min and held until 15.4 min before re-equilibrating to 15% B, held until 20 min. Solvent A was 60:40 MeCN:H_2_O and solvent B was 90:9:1 IPA:MeCN:H_2_O with both solvents containing 0.1% formic acid and 10 mM ammonium formate.

Lipid extracts were transferred to an LC vial and kept in multisampler held at 4°C prior to analysis. For positive ionization, 3 μL samples diluted 30-fold in IPA were injected, whereas 5 μL undiluted sample was run in negative ionization. Ion source parameters for positive ionization were as follows: drying gas at 250°C at 12 L/min, nebulizer at 30 psig, and sheath gas at 300°C flowing 11 L/min. For negative mode, drying gas was the same, nebulizer operated at 35 psig and sheath gas at 375°C flowing 12 L/min. The capillary voltage was 4000 V, skimmer at 75 V, fragmentor at 190 V and octopole RF at 750 V for both ionizations. Reference masses 121.05 and 922.00 m/z were continuously infused during positive ionization whereas 112.98 and 966.00 m/z were used for negative mode. Tandem MS with collision energy at 25 V was performed using iterative MS/MS on six pooled lipid extracts for each ionization. Individual samples were run using MS1 scanning from 100-1500 m/z in each ionization as separate experiments. Agilent LipidAnnotator software was used to create lipid libraries using the MS/MS data and applied during Profinder (v8.0) processing of MS1 data. Further data processing was performed in R.

### Oxylipid extraction

Oxylipids were extracted from 250 μL of freshly thawed plasma (55). To plasma, 1 mL methanol (0.02% BHT) and 5 μL deuterated internal standards at a concentration of 0.30 ng/μL were added (**STable 4**). Samples were vortexed and incubated on ice for 30 mins. After being centrifuged at 4°C for 10 min at 12,000 RPM, clear solvent was transferred to 15 mL conical tubes. Remaining sample pellets were re-extracted with 500 μL MeOH, pooled with the initial 1 mL extract, and dried under nitrogen until the volume was below 500 μL. 6 mL acidified H_2_O (pH=3.5) was added to samples, which were then loaded on SPE columns preconditioned with 6 mL MeOH, then 6 mL H_2_O (pH=7.0). Conical tubes were washed with an additional 2 mL of water (pH=7.0) and loaded onto columns, which were washed with 3 mL hexane. After 10 min of drying, oxylipids were eluted with 4 mL of methyl formate followed by 2 mL of MeOH into conical tubes containing 6 μL MeOH (30% glycerol). Extracts were dried under nitrogen, then tubes washed with 2×400 μL MeOH, transferred to microcentrifuge tubes, then dried again under nitrogen. Oxylipid extracts were reconstituted in 100 μL 1:1 MeOH:H_2_O, transferred to LC vials with 250 μL spring inserts, and stored at -80°C until analysis.

### Targeted oxylipid analysis

LC-MS analysis of oxylipids was performed on an Agilent 1290 Infinity II LC coupled to an Agilent 6495C triple-quadrupole MS which was operated in dynamic multiple reaction monitoring (dMRM) mode. Samples were kept in a multisampler at 4°C for no more than 24h prior to injection. Injection volume was 10 μL. Oxylipids were separated on an Agilent RRHD Eclipse Plus C18 column (2.1×150 mm, 1.8 μm) with an Agilent Eclipse Plus C18 guard (2.1×5 mm, 1.8 μm) that was kept at 40°C. The LC flow was kept constant at 0.350 mL/min with the following gradient: start at 15% B to 33% at 3.5min, to 38% B at 5.5 min, to 42% B at 7 min, to 48% B at 9 min, to 65% B at 15 min, to 75% B at 17 min, to 85% B at 18.5 min, to 95% B at 19.5 min, and finally to 15% B at 21 min, held until 26 min. Solvent A was 0.01% acetic acid and B was 90:10 MeCN:IPA. A blank solvent injection was run between each sample injections.

A list of oxylipids with retention times (RTs) and MRMs is provided as supplement (**STable 4**). RTs were determined using pure standards purchased from Cayman Chemical and compared to published data (56, 57). RTs were predicted for oxylipids without pure standard using regressions of known standard RTs **(STable 4)**. Pooled plasma extracts and standards were used to evaluate the method and adjust scanning windows, which varied from 0.35-0.6 min for each compound. Transitions and MS parameters were adopted from Strassburg *et al*. as follows: drying gas at 290°C at 10 L/min, nebulizer at 35 psi, sheath gas at 350°C at 11 L/min, capillary voltage at 3500 V and nozzle voltage at 1000 V (41). Data was collected and analyzed using Agilent MassHunter Suite. Collected data was manually curated to ensure the correct peaks were picked for target compounds. Compounds present in at least 80% of samples were reported in the final dataset.

### Data analysis

All data was processed in *R* versions 4.1.1 - 4.2.0 (58). For clinical data, the median and quartiles 1 and 3 were reported. Lipidomics data are reported in molar units or pg lipid/mL plasma for oxylipids. These values are ‘semi-quantitative’ because IS and standard curves were not run for every compound analyzed (see ‘Reagents’).

Mean +/- SEM was reported unless otherwise stated. All *P*-values were considered significant if *P*<0.05. For multiple comparisons, *P*-values were adjusted using FDR correction. Due to the exploratory nature of the study, a relaxed *q*<0.30 was considered as significant in our volcano plot analyses. For multiple linear regressions, a stricter *q*<0.15 was used to identify predictors of systolic BP. Correlation plots of were made using the *corrplot* package and Spearman coefficients were reported (59). For linear regressions, independent variables were log_2_ transformed, mean-centered and scaled to the standard deviation prior to analysis. The lasso machine learning model was performed using the *tidymodels* package (60). Data from MIDUS was randomly split into a training and test datasets at a 70/30 ratio. A 5-fold cross validation loop repeated 10 times was used to optimize the model penalty parameter based on the lowest root mean square error value. The final lasso model was then used to predict the training data in entirety, and the unseen test data. For oxylipid-to-fatty acid ratios, we only analyzed LA, ARA, EPA and DHA derived oxylipids because fatty acid precursors for other oxylipids were not measured.

## Supplementary Materials

**STable 1. Results of volcano plot analysis of untargeted analysis in SHOW and MIDUS**.

**STable 2. Results from multiple linear regression for SHOW and MIDUS**.

**STable 3. Oxylipid abbreviations, major enzyme pathways, and volcano plot analyses**.

**STable 4. Details on the oxylipid analysis are provided, including retentions times, transitions, and internal standards to which analytes were normalized**.

**SFigure 1:**
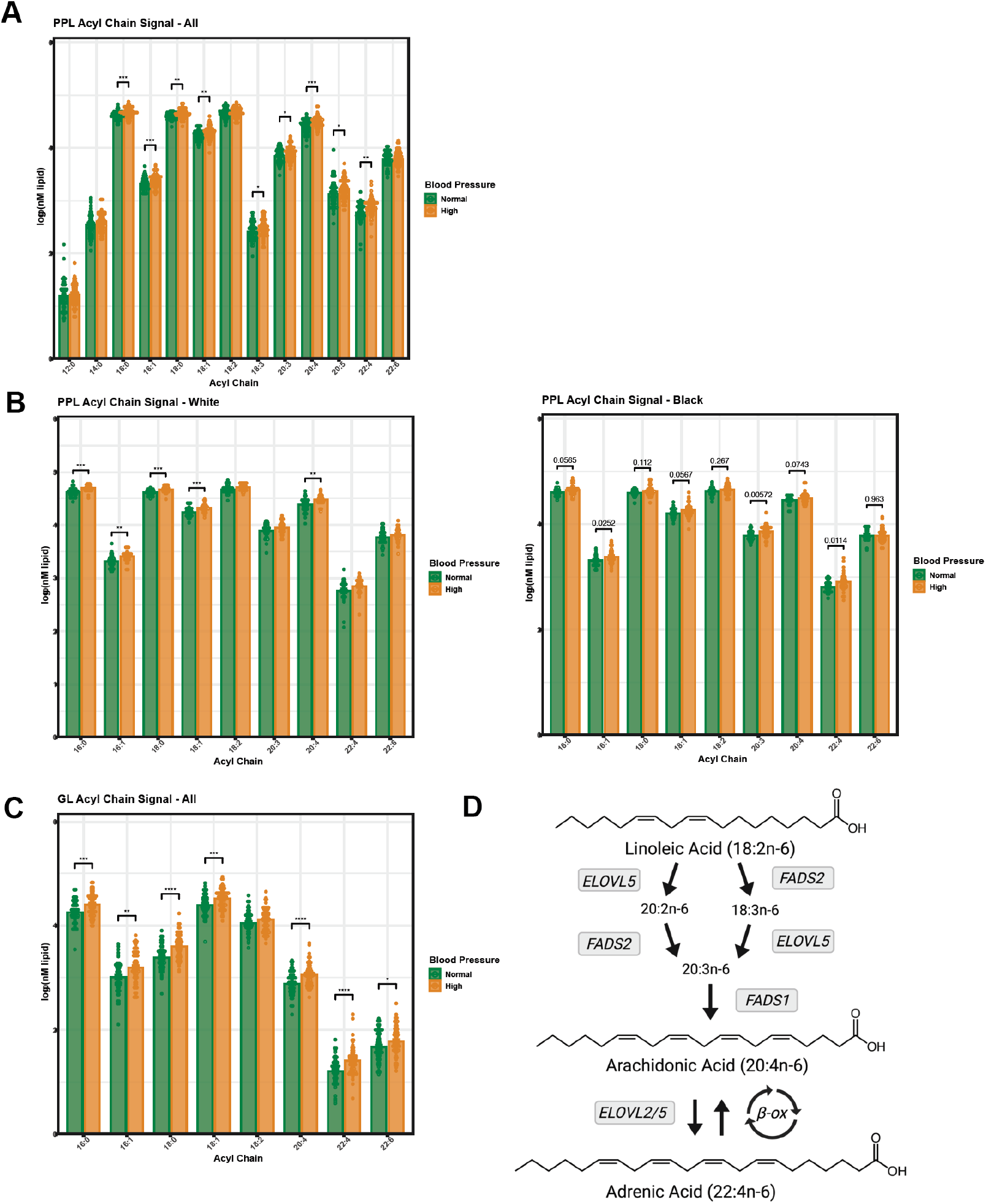
Acyl chain abundance in select lipids classes from the SHOW study. A) An extended distribution of acyl chains detected in the phospholipid (PPL) pool in the SHOW cohort is compared between women with normal blood pressure (Normal) and high blood pressure (High). B) Major acyl chain signal changes from the PPL pool in women with high versus normal BP stratified by race. C) The acyl chain distribution of non-PPL glycerolipids (GL) for the SHOW population, stratified by BP status. D) The arachidonic acid synthesis pathway is provided.

**SFigure 2:**
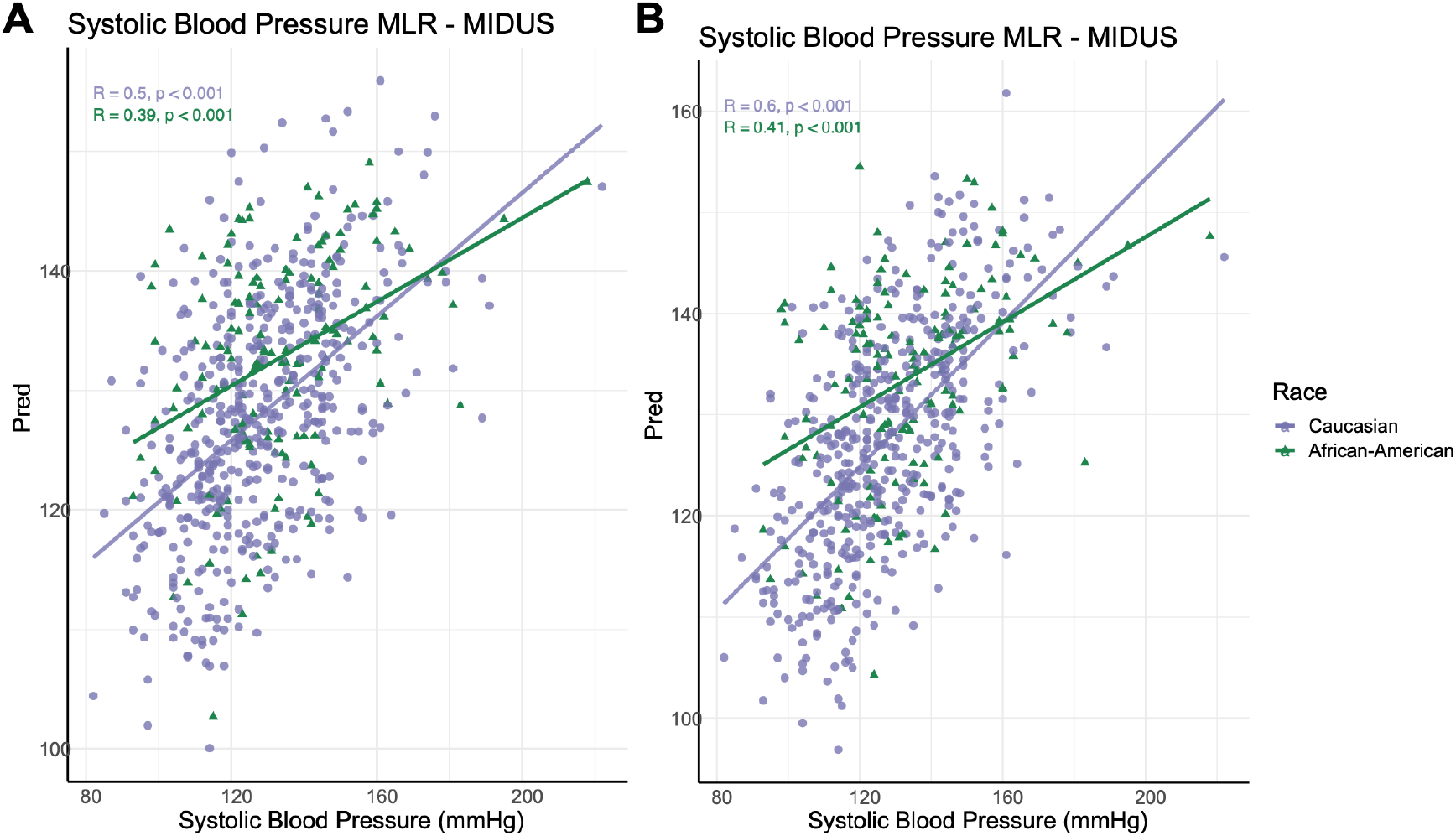
MIDUS multiple linear regression results using all significant predictors containing 18:2, 20:4 or 22:4. A) Systolic blood pressure (BP) predictions compared to actual values from a regression containing all lipids with 18:2, 20:4, and 22:4 that were significantly predictive of systolic BP. B) The covariates age, waist circumference, hemoglobin A1C, and c reactive protein were added to the regression from A and systolic BP predictions are plotted against the quantified lipids.

**SFigure 3:**
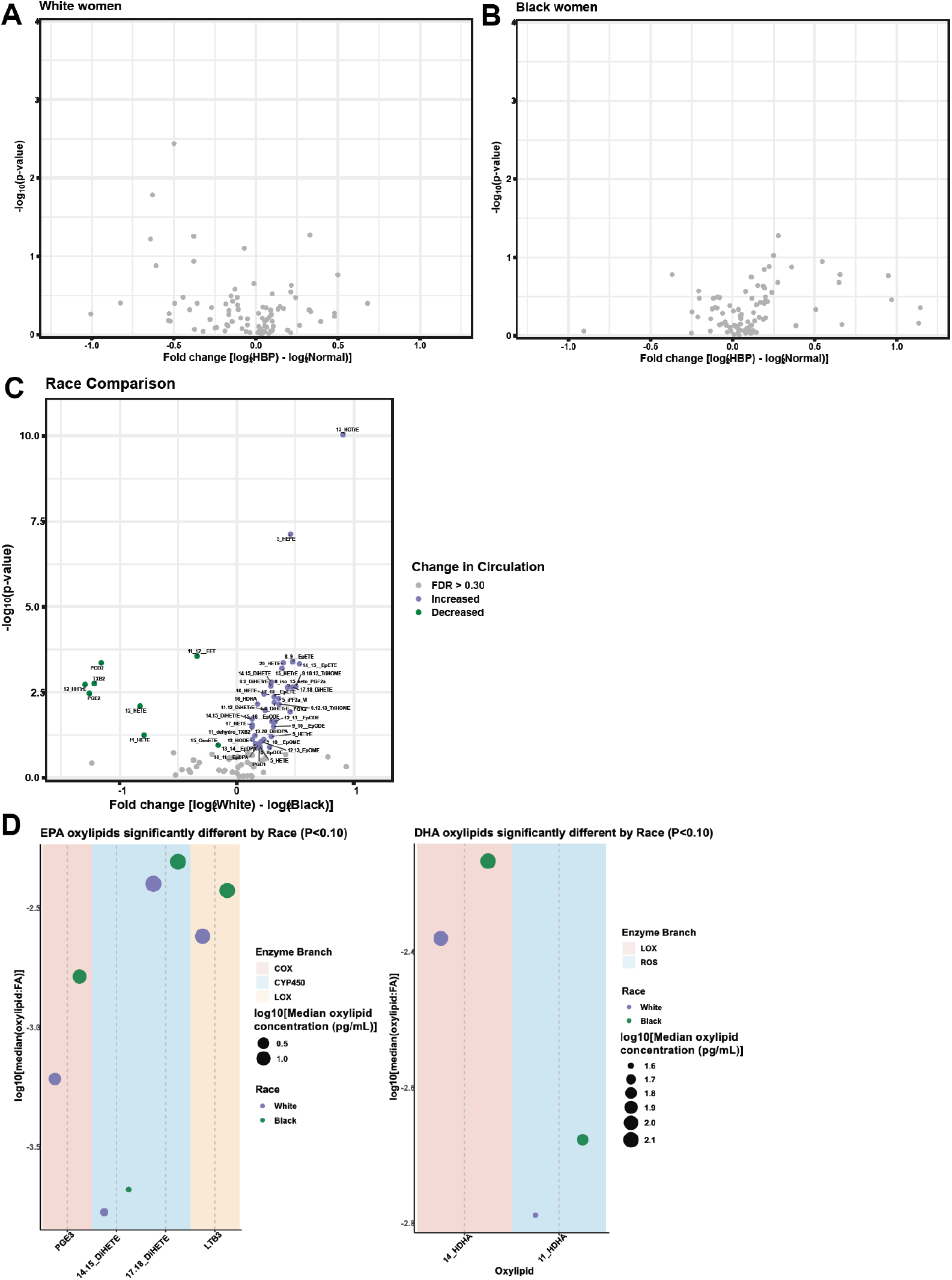
Comparison of oxylipid levels in a subset of the MIDUS cohort. A) Volcano plot of oxylipid changes during HBP in White women after FDR correction. No lipids were significantly different. B) Volcano plot of oxylipid changes during HBP in Black women. C) Volcano plot comparing oxylipid changes between race. D) Median ratio of oxylipid:precursor fatty acid for EPA and DHA derived oxylipids significantly different between races.

## References

1. CDC. Women and Heart Disease. Updated 2020 Accessed May 10, 2022, 2022.

2. Multiple risk factor intervention trial. Risk factor changes and mortality results. Multiple Risk Factor Intervention Trial Research Group. Jama. 1982;248(12):1465–77.

3. Adedinsewo DA, Pollak AW, Phillips SD, Smith TL, Svatikova A, Hayes SN, et al. Cardiovascular Disease Screening in Women: Leveraging Artificial Intelligence and Digital Tools. 2022;130(4):673–90.

4. Keteepe-Arachi T, and Sharma S. Cardiovascular Disease in Women: Understanding Symptoms and Risk Factors. European cardiology. 2017;12(1):10–3.

5. Bybee KA, and Stevens TL. Matters of the heart: cardiovascular disease in U.S. women. Missouri medicine. 2013;110(1):65–70.

6. Mehta LS, Beckie TM, DeVon HA, Grines CL, Krumholz HM, Johnson MN, et al. Acute Myocardial Infarction in Women. 2016;133(9):916–47.

7. Pool LR, Ning H, Lloyd-Jones DM, and Allen NB. Trends in Racial/Ethnic Disparities in Cardiovascular Health Among US Adults From 1999-2012. Journal of the American Heart Association. 2017;6(9).

8. Kalinowski J, Taylor JY, and Spruill TM. Why Are Young Black Women at High Risk for Cardiovascular Disease? Circulation. 2019;139(8):1003–4.

9. Williams RA. Cardiovascular disease in African American women: a health care disparities issue. Journal of the National Medical Association. 2009;101(6):536–40.

10. Zheutlin AR, Caldwell D, Anstey DE, Conroy MB, Ogedegbe O, and Bress AP. Trends in Hypertension Clinical Trials Focused on Interventions Specific for Black Adults: An Analysis of ClinicalTrials.gov. J Am Heart Assoc. 2020;9(24):e018512.

11. Gaillard T, and Osei K. Ethnic differences in serum lipids and lipoproteins in overweight/obese African-American and white American women with pre-diabetes: significance of NMR-derived lipoprotein particle concentrations and sizes. BMJ Open Diabetes Res Care. 2016;4(1):e000246.

12. Ummarino D. Lipidomics refines CVD risk prediction. Nature Reviews Cardiology. 2016;13(12):697-.

13. West AL, Michaelson LV, Miles EA, Haslam RP, Lillycrop KA, Georgescu R, et al. Lipidomic Analysis of Plasma from Healthy Men and Women Shows Phospholipid Class and Molecular Species Differences between Sexes. Lipids. 2021;56(2):229–42.

14. Aristizabal-Henao JJ, Jones CM, Lippa KA, and Bowden JA. Nontargeted lipidomics of novel human plasma reference materials: hypertriglyceridemic, diabetic, and African-American. Analytical and bioanalytical chemistry. 2020;412(27):7373–80.

15. Poss AM, Maschek JA, Cox JE, Hauner BJ, Hopkins PN, Hunt SC, et al. Machine learning reveals serum sphingolipids as cholesterol-independent biomarkers of coronary artery disease. The Journal of clinical investigation. 2020;130(3):1363–76.

16. Mundra PA, Barlow CK, Nestel PJ, Barnes EH, Kirby A, Thompson P, et al. Large-scale plasma lipidomic profiling identifies lipids that predict cardiovascular events in secondary prevention. JCI Insight. 2018;3(17).

17. Gladine C, and Fedorova M. The clinical translation of eicosanoids and other oxylipins, although challenging, should be actively pursued. Journal of mass spectrometry and advances in the clinical lab. 2021;21:27–30.

18. Dyall SC, Balas L, Bazan NG, Brenna JT, Chiang N, da Costa Souza F, et al. Polyunsaturated fatty acids and fatty acid-derived lipid mediators: Recent advances in the understanding of their biosynthesis, structures, and functions. Progress in Lipid Research. 2022;86:101165.

19. Malecki KMC, Nikodemova M, Schultz AA, LeCaire TJ, Bersch AJ, Cadmus-Bertram L, et al. The Survey of the Health of Wisconsin (SHOW) Program: An infrastructure for Advancing Population Health Sciences. medRxiv : the preprint server for health sciences. 2021.

20. Hopkins Tanne J. US guidelines say blood pressure of 120/80 mm Hg is not “normal”. BMJ (Clinical research ed). 2003;326(7399):1104.

21. Fuchs FD, and Whelton PK. High Blood Pressure and Cardiovascular Disease. 2020;75(2):285–92.

22. Tsao CW, Aday AW, Almarzooq ZI, Alonso A, Beaton AZ, Bittencourt MS, et al. Heart Disease and Stroke Statistics-2022 Update: A Report From the American Heart Association. Circulation. 2022;145(8):e153–e639.

23. Flint AJ, Rexrode KM, Hu FB, Glynn RJ, Caspard H, Manson JE, et al. Body mass index, waist circumference, and risk of coronary heart disease: a prospective study among men and women. Obesity research & clinical practice. 2010;4(3):e171–e81.

24. Bentley-Lewis R, Koruda K, and Seely EW. The metabolic syndrome in women. Nature clinical practice Endocrinology & metabolism. 2007;3(10):696–704.

25. Hawkes N. High levels of bad cholesterol in early middle age are linked to CVD risk decades later, study finds. 2019;367:6814.

26. Peters SA, Singhateh Y, Mackay D, Huxley RR, and Woodward M. Total cholesterol as a risk factor for coronary heart disease and stroke in women compared with men: A systematic review and meta-analysis. Atherosclerosis. 2016;248:123–31.

27. Lewington S, Whitlock G, Clarke R, Sherliker P, Emberson J, Halsey J, et al. Blood cholesterol and vascular mortality by age, sex, and blood pressure: a meta-analysis of individual data from 61 prospective studies with 55,000 vascular deaths. Lancet (London, England). 2007;370(9602):1829–39.

28. Kontush A, Lhomme M, and Chapman MJ. Unraveling the complexities of the HDL lipidome. Journal of lipid research. 2013;54(11):2950–63.

29. Radler BT. The Midlife in the United States (MIDUS) Series: A National Longitudinal Study of Health and Well-being. Open health data. 2014;2(1).

30. Vatsa N, Thobani A, Buendia L, Murphy K, Asier S, Chen Z, et al. Cardiovascular Risk Factors in Younger Black Women: Results from the 10,000 Women Community Screening Project. American Heart Journal Plus: Cardiology Research and Practice. 2021;8:100037.

31. Berkowitz L, Henríquez MP, Salazar C, Rojas E, Echeverría G, Love GD, et al. Association between serum sphingolipids and eudaimonic well-being in white U.S. adults. Scientific reports. 2021;11(1):13139.

32. Gabbs M, Leng S, Devassy JG, Monirujjaman M, and Aukema HM. Advances in Our Understanding of Oxylipins Derived from Dietary PUFAs. Advances in nutrition (Bethesda, Md). 2015;6(5):513–40.

33. Ellero-Simatos S, Beitelshees AL, Lewis JP, Yerges-Armstrong LM, Georgiades A, Dane A, et al. Oxylipid Profile of Low-Dose Aspirin Exposure: A Pharmacometabolomics Study. Journal of the American Heart Association. 2015;4(10):e002203.

34. Nayeem MA. Role of oxylipins in cardiovascular diseases. Acta pharmacologica Sinica. 2018;39(7):1142–54.

35. Bai H-W, and Zhu BT. Strong activation of cyclooxygenase I and II catalytic activity by dietary bioflavonoids*. Journal of lipid research. 2008;49(12):2557–70.

36. Bock M, Karber M, and Kuhn H. Ketogenic diets attenuate cyclooxygenase and lipoxygenase gene expression in multiple sclerosis. EBioMedicine. 2018;36:293–303.

37. Mallat Z, Lambeau G, and Tedgui A. Lipoprotein-Associated and Secreted Phospholipases A<sub>2</sub> in Cardiovascular Disease. 2010;122(21):2183–200.

38. Mathias RA, Sergeant S, Ruczinski I, Torgerson DG, Hugenschmidt CE, Kubala M, et al. The impact of FADS genetic variants on ω6 polyunsaturated fatty acid metabolism in African Americans. BMC genetics. 2011;12:50.

39. Rett BS, and Whelan J. Increasing dietary linoleic acid does not increase tissue arachidonic acid content in adults consuming Western-type diets: a systematic review. Nutrition & metabolism. 2011;8:36.

40. Taber L, Chiu CH, and Whelan J. Assessment of the arachidonic acid content in foods commonly consumed in the American diet. Lipids. 1998;33(12):1151–7.

41. Lau ES, Binek A, Parker SJ, Shah SH, Zanni MV, Van Eyk JE, et al. Sexual Dimorphism in Cardiovascular Biomarkers: Clinical and Research Implications. Circulation research. 2022;130(4):578–92.

42. Reiner AP, Beleza S, Franceschini N, Auer PL, Robinson JG, Kooperberg C, et al. Genome-wide association and population genetic analysis of C-reactive protein in African American and Hispanic American women. American journal of human genetics. 2012;91(3):502–12.

43. Thorand B, Löwel H, Schneider A, Kolb H, Meisinger C, Fröhlich M, et al. C-Reactive Protein as a Predictor for Incident Diabetes Mellitus Among Middle-aged Men: Results From the MONICA Augsburg Cohort Study, 1984-1998. Archives of Internal Medicine. 2003;163(1):93–9.

44. Quintana LF, Guzmán B, Collado S, Clària J, and Poch E. A coding polymorphism in the 12-lipoxygenase gene is associated to essential hypertension and urinary 12(S)-HETE. Kidney international. 2006;69(3):526–30.

45. Yeung J, and Holinstat M. 12-lipoxygenase: a potential target for novel anti-platelet therapeutics. Cardiovascular & hematological agents in medicinal chemistry. 2011;9(3):154–64.

46. Fernandez-Jimenez R, Wang TJ, Fuster V, and Blot WJ. Low-Dose Aspirin for Primary Prevention of Cardiovascular Disease: Use Patterns and Impact Across Race and Ethnicity in the Southern Community Cohort Study. Journal of the American Heart Association. 2019;8(24):e013404.

47. Davidson KW, Barry MJ, Mangione CM, Cabana M, Chelmow D, Coker TR, et al. Aspirin Use to Prevent Cardiovascular Disease: US Preventive Services Task Force Recommendation Statement. Jama. 2022;327(16):1577–84.

48. Aggarwal R, Chiu N, Wadhera RK, Moran AE, Raber I, Shen C, et al. Racial/Ethnic Disparities in Hypertension Prevalence, Awareness, Treatment, and Control in the United States, 2013 to 2018. Hypertension (Dallas, Tex : 1979). 2021;78(6):1719–26.

49. Martinez JA, Skiba MB, Chow HS, Chew WM, Saboda K, Lance P, et al. A Protective Role for Arachidonic Acid Metabolites against Advanced Colorectal Adenoma in a Phase III Trial of Selenium. Nutrients. 2021;13(11).

50. Mattmiller SA, Carlson BA, Gandy JC, and Sordillo LM. Reduced macrophage selenoprotein expression alters oxidized lipid metabolite biosynthesis from arachidonic and linoleic acid. The Journal of nutritional biochemistry. 2014;25(6):647–54.

51. Ubhi BK. Direct Infusion-Tandem Mass Spectrometry (DI-MS/MS) Analysis of Complex Lipids in Human Plasma and Serum Using the Lipidyzer™ Platform. Methods in molecular biology (Clifton, NJ). 2018;1730:227–36.

52. Eichelmann F, Sellem L, Wittenbecher C, Jäger S, Kuxhaus O, Prada M, et al. Deep Lipidomics in Human Plasma: Cardiometabolic Disease Risk and Effect of Dietary Fat Modulation. 2022;146(1):21–35.

53. Matyash V, Liebisch G, Kurzchalia TV, Shevchenko A, and Schwudke D. Lipid extraction by methyl-tert-butyl ether for high-throughput lipidomics. J Lipid Res. 2008;49(5):1137–46.

54. Jain R, Wade G, Ong I, Chaurasia B, and Simcox J. Determination of tissue contributions to the circulating lipid pool in cold exposure via systematic assessment of lipid profiles. Journal of lipid research. 2022;63(7):100197.

55. Ostermann AI, Willenberg I, and Schebb NH. Comparison of sample preparation methods for the quantitative analysis of eicosanoids and other oxylipins in plasma by means of LC-MS/MS. Analytical and bioanalytical chemistry. 2015;407(5):1403–14.

56. Strassburg K, Huijbrechts AM, Kortekaas KA, Lindeman JH, Pedersen TL, Dane A, et al. Quantitative profiling of oxylipins through comprehensive LC-MS/MS analysis: application in cardiac surgery. Analytical and bioanalytical chemistry. 2012;404(5):1413–26.

57. Misheva M, Kotzamanis K, Davies LC, Tyrrell VJ, Rodrigues PRS, Benavides GA, et al. Oxylipin metabolism is controlled by mitochondrial β-oxidation during bacterial inflammation. Nature Communications. 2022;13(1):139.

58. Team RC. R: A language and environment for statistical computing. R Foundation for Statistical Computing. https://www.R-project.org/. 2022.

59. Taiyun Wei VS, Michael Levy, Yihui Xie, Yan Jin, Jeff Zemla, Moritz Freidank, Jun Cai, Tomas Protivinsky. Visualization of a Correlation Matrix. https://github.com/taiyun/corrplot.

60. Kuhn M, Wickham, Hadley. Tidymodels: a collection of packages for modeling and machine learning using tidyverse principle. 2020.

